# Standardizing annual dengue intensity reveals global drivers of transmission

**DOI:** 10.64898/2025.12.19.25342670

**Authors:** Abigail J. Porzucek, Rafael Lopes, Yi Ting Chew, Ke Li, Oliver J. Brady, Joshua L. Warren, Colin J. Carlson, Daniel M. Weinberger, Nathan D. Grubaugh

## Abstract

Dengue incidence is increasing due to climate change and globalization, as well as enhanced case ascertainment. Despite the increasing transmission intensity of dengue virus, there is no standardized measure of disease burden, nor a standardized outbreak definition. Without such measures, it is difficult to understand how transmission patterns are changing over time and to identify what drives these changes globally. To address these problems, we developed a globally standardized metric of dengue burden, which we refer to as the Relative Intensity Score (RISc). RISc is an annual, country-level metric that measures short-term aberrations in dengue incidence relative to location- and time-specific baselines. We derived RISc estimates for 57 countries from 1990-2023 and then used RISc to identify years in which each country experienced outbreak-level transmission. Using these measures, we then examined patterns of RISc over time and found that years with increased RISc tend to cluster together, suggesting that changes in intensity are sustained through multi-year cycles. Finally, we identified factors associated with increased RISc on global and regional scales and found that increases in temperature were associated with the greatest increases in intensity. In sum, this study provides a globally standardized measure of dengue burden and demonstrates how this metric can be utilized to uniformly identify outbreak occurrence, track changing epidemiologic patterns, and identify environmental factors driving changes in intensity.

## Introduction

Dengue is the most prevalent mosquito-borne viral disease, with roughly half the world population currently at risk. Over the past several decades, a steady rise in dengue incidence has been observed, with this trend expected to continue into the future.^1–3^ Yet, it remains unclear how incidence reported in recent years compares to incidence in past decades, given that the baseline transmission dynamics as well as case definitions, reporting requirements, and access to diagnostics have all changed over the same time frame.^4–6^ This concurrent increase in transmission and case ascertainment makes it difficult to untangle short-term fluctuations in incidence from long-term changes to baseline levels of dengue.

There are several known drivers that contribute to increases in dengue burden. First, climate change is estimated to account for nearly 20% of dengue cases, with that percentage expected to increase under future climate change scenarios.^7^ Cycles of the El Niño Southern Oscillation (ENSO) influence temperature, rainfall, and humidity, leading to changes in dengue virus transmission, with increases in transmission often observed during hotter and drier El Niño phases of ENSO.^8,9^ Further, climate change has expanded the geographic areas suitable to the vectors of dengue virus, *Aedes aegypti* and *Aedes albopictus*.^10,11^ Over many years, this has resulted in northern spread of dengue in the Northern Hemisphere, southern spread of dengue in the Southern Hemisphere, and spread to increased elevation globally.^11^ Additionally, changes in public health response to dengue have influenced the proportion of infections that are detected and reported. As the burden of dengue has increased, so too have surveillance and diagnostic efforts leading to increased detection and reporting of cases.^2,4,12^

Prior efforts to provide global standardized measures of dengue burden have largely focused on deriving force of infection estimates.^13–19^ These methods require serologic data, which measure the fraction of the population that has been exposed, but typically cannot provide insights into the timings of those exposures. As such, these estimates are mostly static and describe how transmission intensity varies country-to-country, but not over time.^13,15,20^ More recent efforts have derived annual force of infection estimates; however, these methods rely on age-stratified case data which have limited availability, and as such have only been conducted in a select few countries, rather than at a global scale.^16–18^ Other options to quantify dengue intensity include more rudimentary measures, such as incidence rates, that do not provide a full picture of changing epidemiologic patterns. Even modest changes in incidence rates are significant in settings with a low baseline for dengue, whereas similar changes in incidence rates would be unremarkable in locations with high baseline burden. Furthermore, there are no standardized dengue outbreak definitions.^5^ Incidence thresholds for outbreaks are often determined locally and can vary considerably among countries and time periods. While this approach is effective in triggering necessary public health responses, it is not conducive to studying factors driving outbreaks on global levels. Thus, we need a uniform, time-varying measure of dengue burden and outbreak occurrence that can be derived from routinely collected data.

To address these issues, we developed a standardized metric of dengue burden that captures annual deviations of reported cases from location- and time-specific baselines. To do this, we compiled more than three decades of surveillance data from 57 countries in 5 distinct global regions. We then modeled long-term trends in incidence at the national level and measured intensity as annual aberrations of reported incidence relative to these trends. As a result, we compared relative short-term variations in dengue intensity across years and locations. We further classified our continuous measure of intensity into three categories: low intensity, expected intensity, and high intensity, with high intensity representing outbreak-like conditions. Finally, we identified that climate factors, specifically elevated temperatures, are correlated with increased short-term dengue burden. In sum, our study provides (1) a retrospective globally standardized measure of transmission intensity; (2) identification of times and places with outbreak-level transmission; and (3) a description of factors associated with increased transmission intensity.

## Results

### Identifying data ambiguities and trends in incidence

To analyze national dengue patterns over several decades and across regional and global scales, we first assessed the completeness of several sources of dengue surveillance data including repositories from OpenDengue, the Pan-American Health Organization (PAHO), World Health Organization (WHO), Gideon Dengue Global Status Report, and Asian Dengue Voice and Action (ADVA). OpenDengue was the most complete single source of data and was used to obtain all case data used in this analysis.^1,2,21–23^ For countries in the Northern Hemisphere, where the “dengue season” peaks in around mid-year, we used the annual case counts.^24^ However, in the Southern Hemisphere, peak dengue virus transmission tends to extend through December and January. Therefore, we used monthly data (when available) from Southern Hemisphere countries to define annual cases from July to June the following calendar year (e.g., for these countries, the 2023-24 transmission season is listed as 2023). To remove countries with inconsistent reporting, we included countries if they had either (1) a minimum of 500 reported annual dengue cases on average between 1990-2023; or (2) a minimum of 10 annual cases per 100,000 population on average during the same time period. We excluded any locations with missing data or zero reported cases accounting for more than 15 years, as well as locations that did not have any population data available through the World Bank.^25^ Starting from a list of 129 countries with reported dengue cases (**Figure S1**), this resulted in a dataset encompassing 57 countries over a 34-year period with 1,995 observations of dengue case data, and 43 country-year combinations with no reported data (**Figure 1a**). Additionally, we calculated the dengue incidence per 100,000 population for each year and country and found a general trend in increasing incidence rates over the 34 years (**Figure 1b**). Yet, even after applying these criteria, additional methods are required to compare dengue incidence across large spans of space and time.

**Figure 1.**
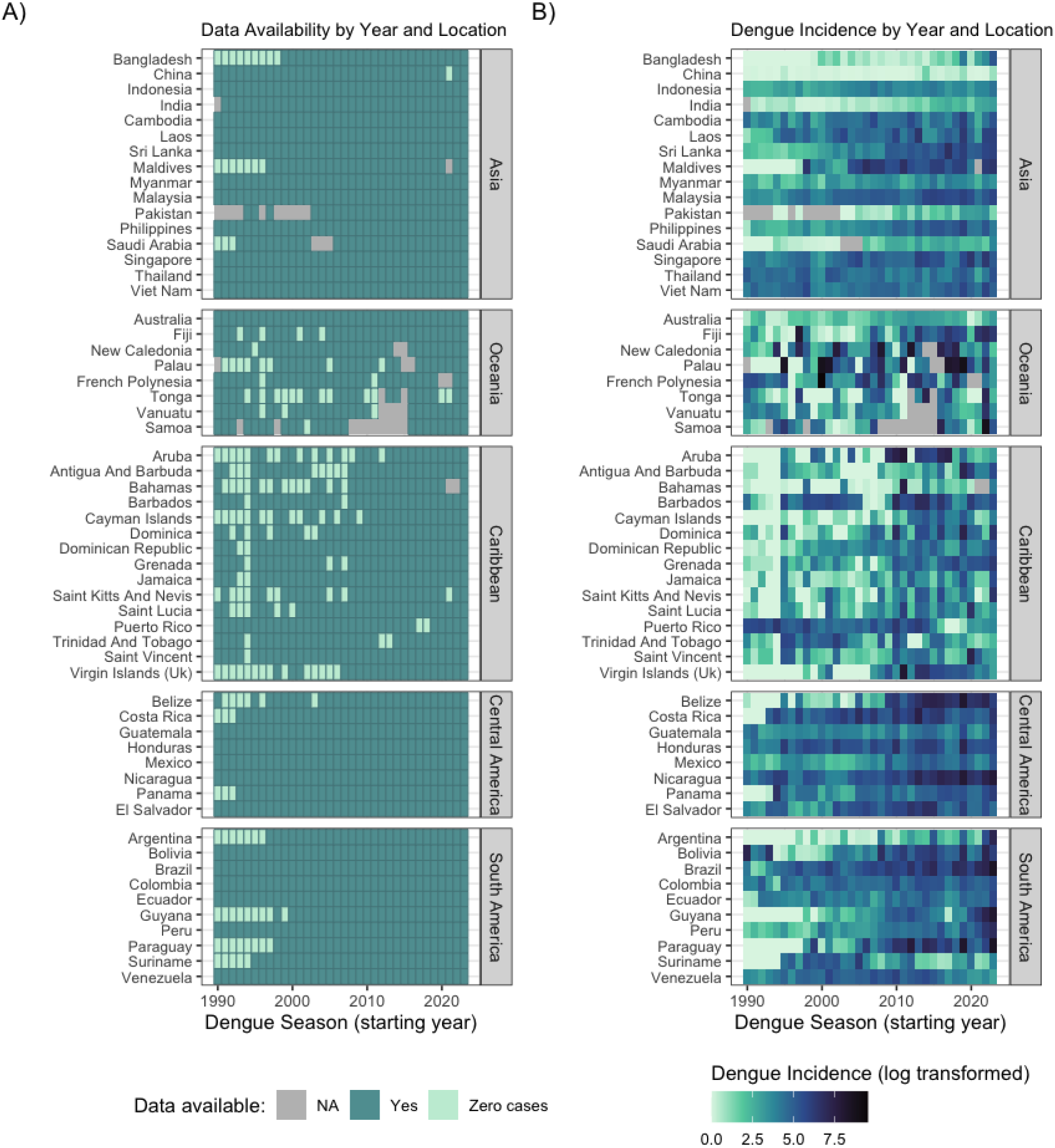
Heterogeneous availability of dengue data and increasing incidence trends. (**A**) Locations and timepoints included in this analysis. Blue tiles show locations and years where at least 1 case was reported. Light green tiles show locations and years where zero cases were reported. Grey tiles represent instances of no reporting. (**B**) Dengue incidence per 100,000 population by country and year. Grey tiles show unavailable data. Dengue case data obtained from OpenDengue.

### Extracting long-term trends to quantify and categorize dengue burden

Dengue incidence has been increasing globally over the past several decades (**Figure 1b**). As the dengue burden has increased, variability in surveillance activities has made it difficult to compare cases across space and time.^2,4,12^ To address this problem, we built a hierarchical Bayesian Poisson regression model to disentangle short-term variations in dengue incidence from long-term trends and to identify factors associated with short-term variations in transmission intensity (**Figure 2**), thereby allowing us to compare annual dengue intensity across 57 countries for more than 30 years.

**Figure 2.**
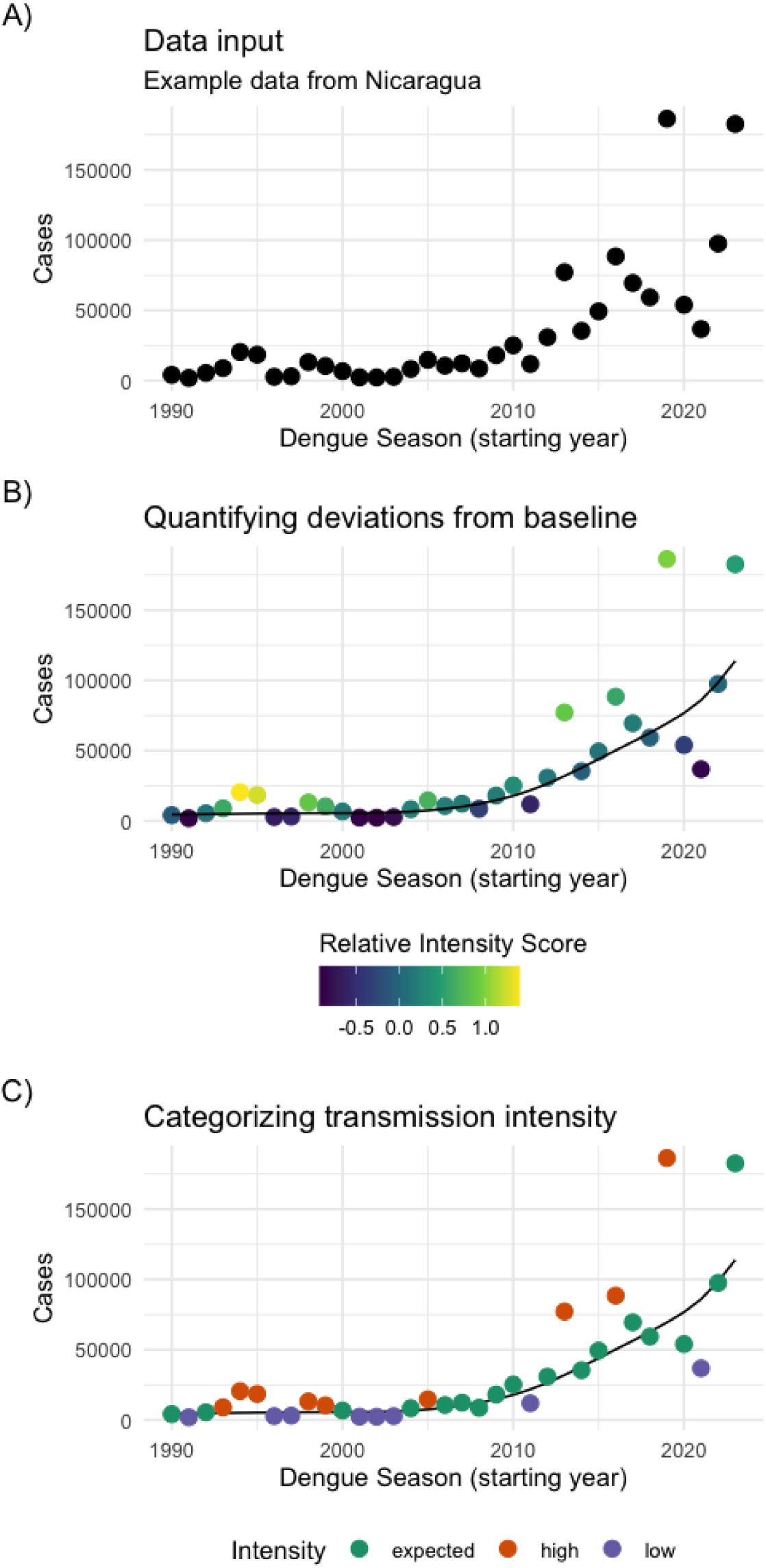
Extracting long-term dengue trends to quantify relative intensity. All example data shown are from Nicaragua. (**A**) Time series case data were included as input into the model. (**B**) Model output decomposed into short-term fluctuations (Relative Intensity Score) and long-term trend. (**C**) RISc is grouped into classes of high intensity, low intensity, and intensity close to baseline levels.

Using the annual reported dengue incidence as the input (**Figure 2a**) and controlling for population size, we used a mixture of smoothing functions and random effects to model dengue burden (**Figure 2b**). Second order random walk smoothing functions were used to model the long-term trends of the incidence data. Log-transformed random effects were used to quantify annual aberrations in incidence in relation to those long-term trends, and we derived Relative Intensity Scores (RISc) from the sum of the random effects. Our RISc metric provides a quantification of dengue burden for each country and year in our dataset, relative to time- and location-specific contexts (**Figure 2c and Figures S2-S6**). Positive RISc represents incidence above baseline and negative RISc represents incidence below baseline, with baseline being equal to a RISc of 0. Our model was built to capture global, continent-level, and country-level random effects. These random effects help account for inconsistencies in surveillance systems by adjusting for unmeasured, location-specific, and time-varying biases in case reporting as well as true long-term increases in incidence over time. Finally, we converted the continuous RISc measure into three categories: low intensity (RISc < -0.5), which represents years in which reported cases were at least 40% lower than expected; intensity within an expected incidence range (RISc -0.5 to 0.5); and high intensity (RISc ≥ 0.5), which is representative of outbreak-like conditions and represents years where reported cases were at least 60% higher than expected (**Figure 2d**). The cutoff point of RISc +/-0.5 was used as it balanced sensitivity and specificity of high and low intensity events. To understand how this compared to outbreaks identified in practice, we compared our threshold to outbreaks of global importance reported in the WHO Disease Outbreak News since 2005 as well as those described in literature (**Figure S7**).^2,26^ We found that our cutoff value of RISc ≥ 0.5 included 5 of the 6 outbreaks identified by the WHO in this period, and captured 14 of the 23 outbreaks described in literature.

After conducting this analysis simultaneously for all 57 countries, we identified RISc ranging from - 6.28 (China, 2021) to 7.09 (Fiji, 1997), which represents reported cases that were roughly 1,000 times higher than expected (**Figure 3a**). We then identified 600 instances of low intensity transmission, 772 instances of transmission within the expected range, and 566 instances of high intensity transmission (**Figure 3b**). Additionally, we imputed gaps in case reporting (**Figure 1a**) based on the estimated baseline for the year of missing data and further informed by the magnitude of global and continental random effects identified in that year (**Figure S8**). Overall, we developed a metric (RISc) that normalizes dengue incidence across locations and time points (**Figure 3a**), which is not possible when using the raw incidence rates alone (**Figure 1b**). This allows us to use RISc to understand what drives changes in dengue intensity, despite differences that may occur from factors such as location-specific case definitions or time-specific reporting practices.

**Figure. 3.**
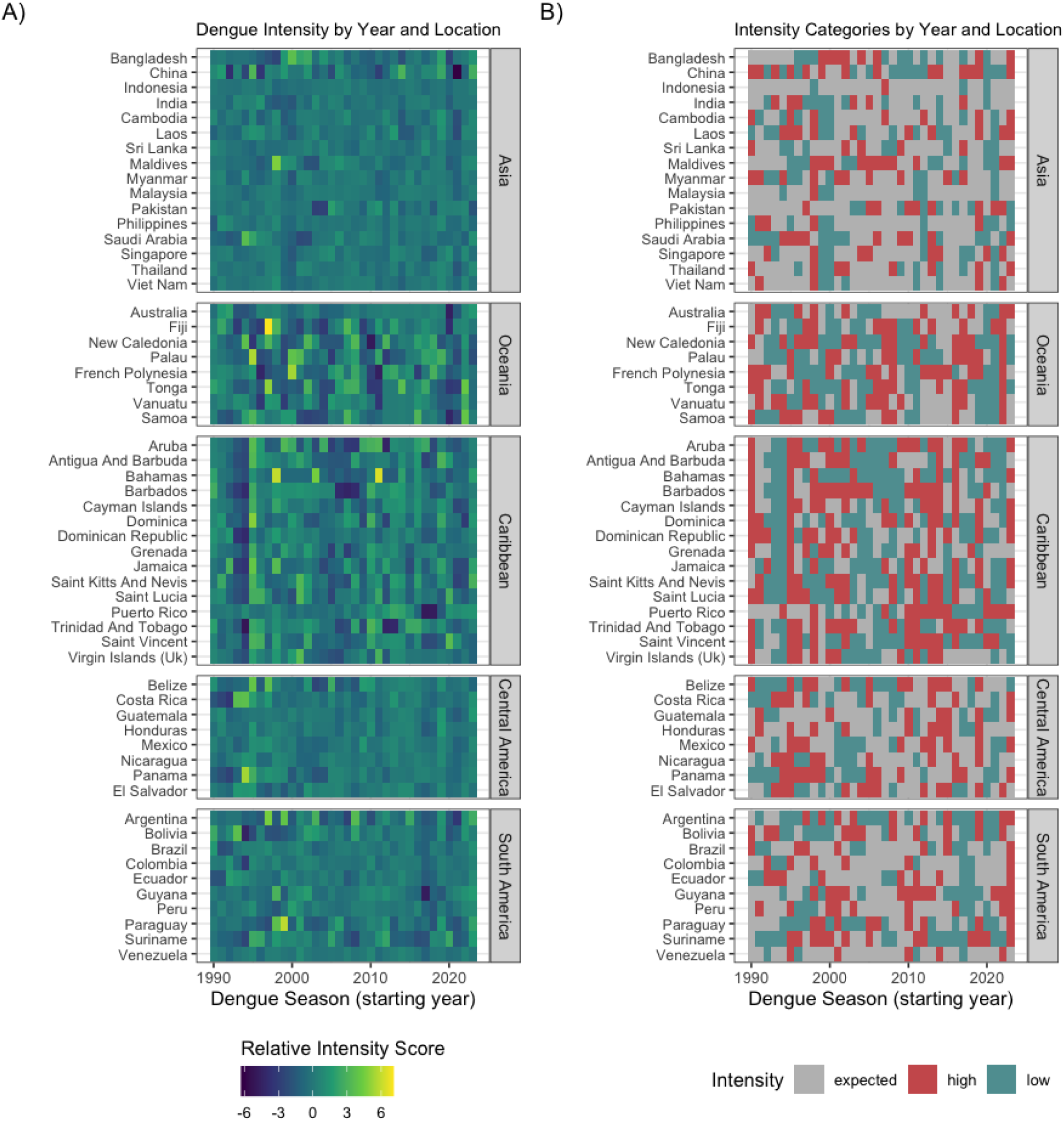
Relative dengue intensity modeled for 57 countries over more than 30 years. (**A**) RISc for each location and time point. (**B**) Intensity categories for each location and time point. RISc >= 0.5 represents higher than expected incidence, and outbreak-like conditions; RISc <= -0.5 represents lower than expected incidence; RISc between those points represents incidence near baseline expectations.

### Exploring global trends in dengue intensity

After establishing our RISc metric, we used it to examine how global and regional intensity varied across time (**Figure 4**). Specifically, we examined if there were time-dependent patterns to intensity and if there was regional or global synchrony amongst high intensity years. To do this, we first extracted global and region-level RISc for each transmission season (**Figure 4a**). We found that years with positive RISc values tended to cluster together in multi-year groups, as did years with negative RISc values. Globally, while many years had RISc values within the range of expected transmission (27 years), we also identified 8 years with global aberrations spread throughout the time series. Specifically, we identified 4 years with low intensity transmission (global RISc ≤ -0.5) and 4 years with high intensity transmission (global RISc ≥ 0.5). The years with the most extreme RISc occurred in 1995 (RISc = 1.11), 1998 (RISc = 0.59), 2023 (RISc = 0.55), and 2021 (RISc = -1.01). On a regional level, we found that all regions had positive RISc values in 1995, with the Caribbean (RISc = 2.62) and Central America (RISc = 1.80) contributing most to the global increase in RISc. In 1998, our regional analysis again showed that all regions had positive RISc, with the Caribbean (RISc = 1.12) and Oceania (RISc = 0.53) contributing most to the global burden. In 2023, four of the five regions had positive RISc, with South America (RISc = 1.22), Central America (RISc = 0.77), and the Caribbean (RISc = 0.50) contributing most to the global increase. However, in 2021, all regions had negative RISc values, which were likely associated with surveillance or transmission disruptions during the COVID-19 pandemic.

**Figure 4.**
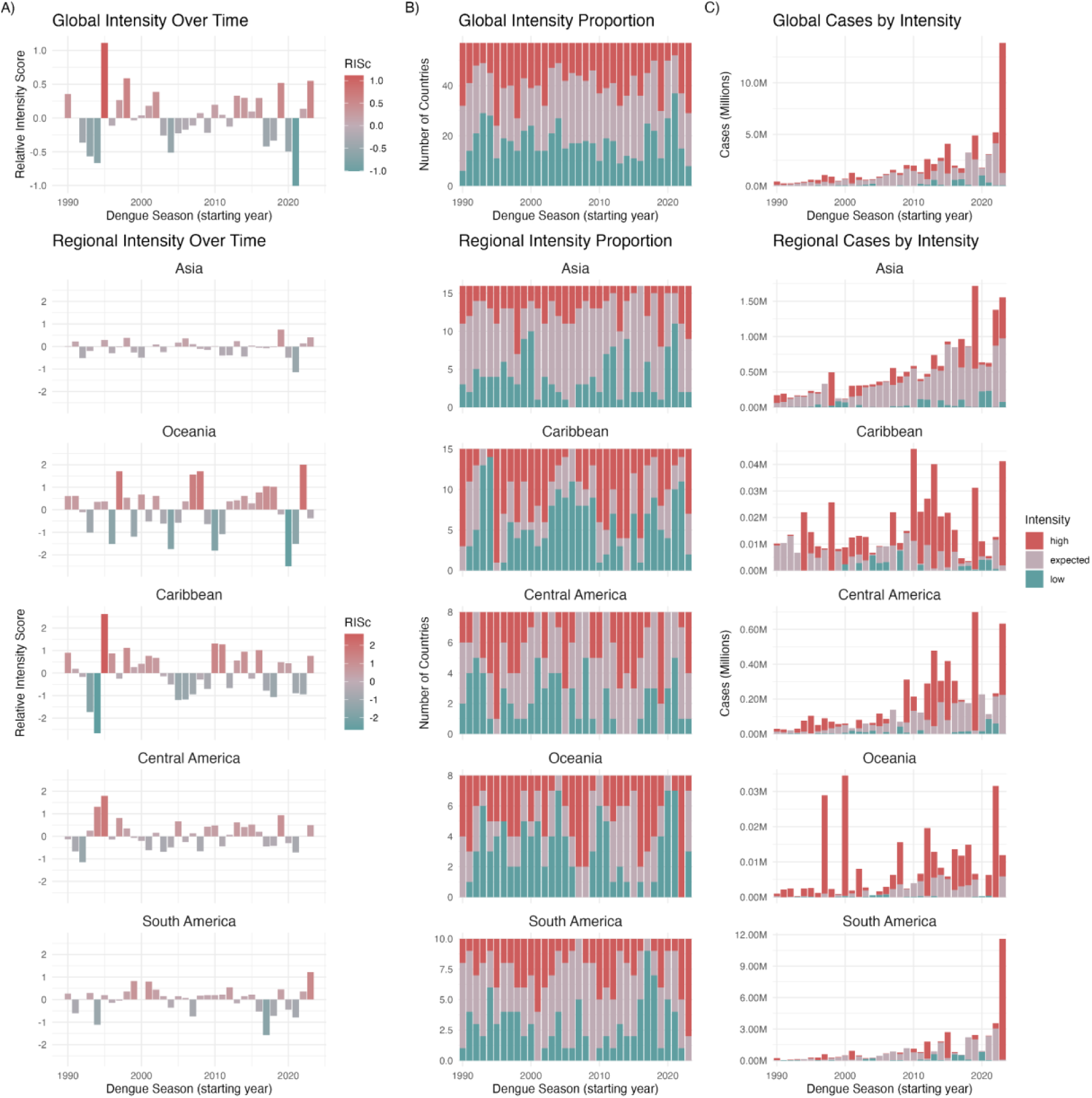
Trends in global and regional dengue intensity. (**A**) Annual country-level RISc averaged over global and regional scales. (**B**) The number of countries globally and regionally experiencing high, expected, and low transmission intensity each year. (**C**) The number of total cases coming from high, expected, and low transmission settings globally and regionally.

We then examined how the patterns of annual intensity contributed to RISc fluctuations by extracting the number of countries experiencing low, expected, or high transmission intensity regionally and globally (**Figure 4b**). As expected, we found that globally, the years with the highest RISc values also have the most countries experiencing high intensity transmission, with 33 countries having high RISc in 1995, 30 countries having high RISc in 1998, and 29 countries having high RISc in 2019. Regionally, we found that some regions have greater co-occurrence among high intensity years and wider dispersion of RISc than others (**Figure S9**). Oceania had the highest degree of co-occurrence, with all 9 countries in the region (100%) experiencing high intensity transmission in 2022. In contrast, Asia had the lowest variation in RISc and typically had the highest proportion of countries within the “expected” range, with 14 countries (88%) having expected intensity in 2016, the year in which Asia had the fewest countries experiencing co-occurring high-intensity seasons.

Finally, we calculated the number of cases coming from high, expected, and low intensity settings each year (**Figure 4c**). Overall, we found that cases from high RISc settings represented a disproportionate amount of total reported cases. By far, the 2023 season (which extended into 2024 for the Southern Hemisphere countries) had the most cases (12,590,818) coming from high intensity countries, representing 90.9% of all reported cases. This was primarily driven by Brazil, which reported 10,047,185 cases in 2023-24, which was 79.8% of all high-intensity cases reported that year.

### Identifying large-scale moderators of dengue intensity

Finally, we identified correlates of annual dengue intensity. To do this, we conducted a meta-regression analysis on RISc values to examine the association of climate, socioeconomic, and time-related variables with RISc globally and regionally (**Figure 5 and Figure S10**). RISc values were estimated from the first stage model using posterior means, and posterior standard deviations were used to characterize uncertainty in the estimates during the second stage model. We selected covariates that had temporal variation, could be measured on annual and national scales, and that prior research had shown to influence dengue dynamics.^27–36^ In particular, we used temperature and precipitation anomalies to measure large-scale variations in climate patterns. On a global scale, temperature anomalies were associated with the greatest increase in RISc, with higher anomalies leading to higher RISc (0.21, CI: 0.13, 0.30; **Figure 5a**). However, precipitation anomalies were not associated with RISc (-0.05, CI: -0.11, 0.02). We hypothesized that population immunity would also impact RISc. However, in the absence of seroprevalence studies, we used the previous year’s RISc as these values would likely be correlated with changing population immunity. We found positive RISc values from the prior year were associated with higher RISc values (0.17, CI: 0.10, 0.24), suggesting that high-intensity transmission extends through multiple seasons (**Figure 4a**). Additionally, our analysis showed that more recent years were associated with lower RISc (-0.28, CI: -0.38, -0.18), indicating that the largest RISc values occurred during the earlier years of our dataset. All other variables we included in our analysis, namely urbanicity, gross domestic product (GDP), forest area, and access to electricity, were not significantly associated with RISc. Regional correlations with RISc were largely in line with global results. Regionally, we found that temperature anomalies were associated with increased RISc in all regions except for Oceania. Likewise, we found that elevated RISc in the previous year was associated with elevated RISc the following year in all regions.

**Figure 5.**
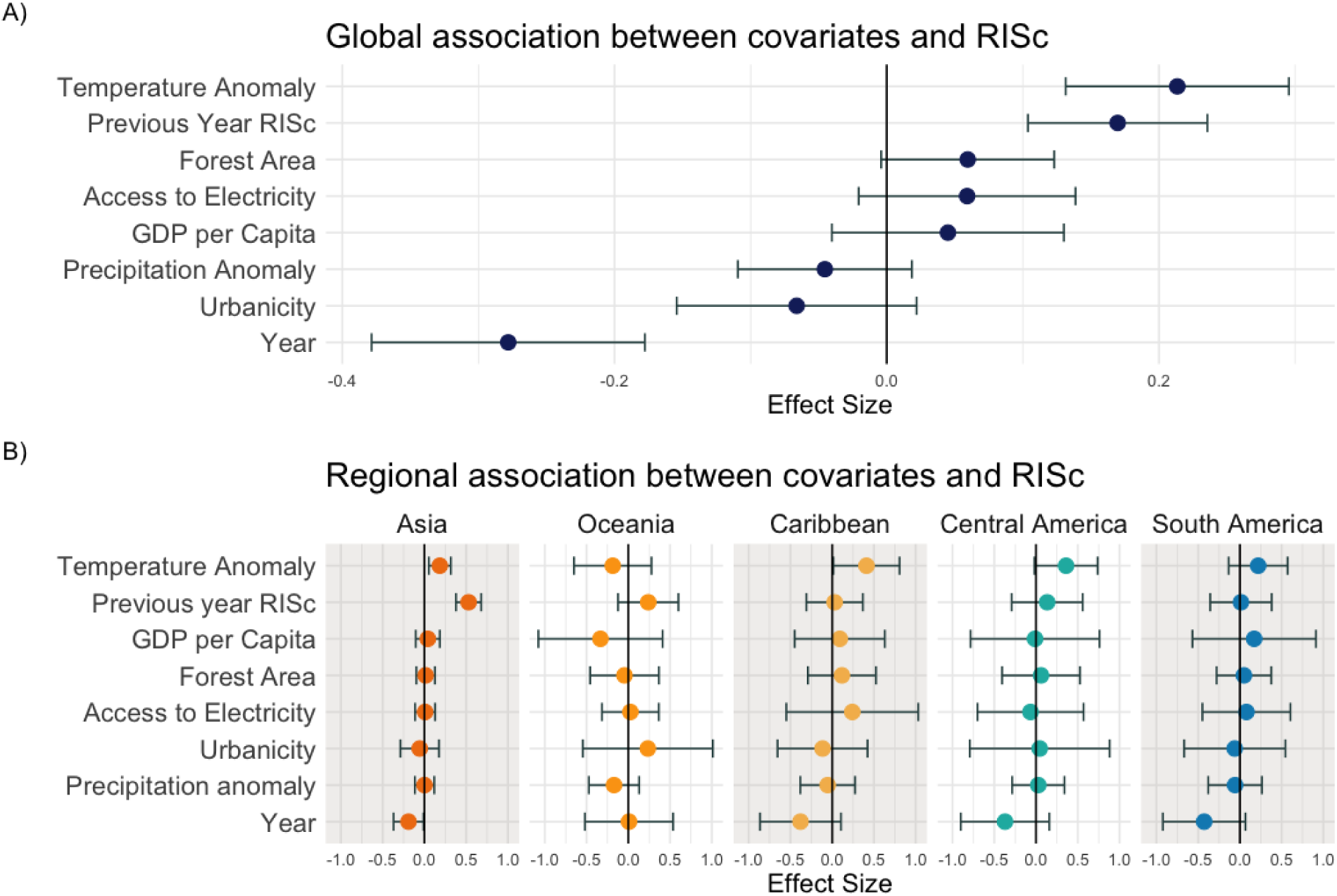
Results of the meta-regression analysis to identify drivers of global and regional dengue intensity. Effect sizes greater than 0 show positive association with RISc and effect sizes less than 0 show negative association with RISc. Effect size point estimates are shown in colored dots and error bars show 95% confidence intervals. (**A**) Impacts of external drivers on a global scale. (**B**) Impacts of external drivers on regional scales.

In sum, we identified that temperature anomalies had the greatest positive impact on RISc, with hotter temperatures leading to higher RISc values. We also found that measures related to population immunity, namely the previous year’s intensity, had a significant impact on RISc and suggest that high intensity years tend to be sequential.

## Discussion

Dengue cases have continued to increase over the past several decades.^1,2^ This upward trend is likely due to true increases in transmission as well as better surveillance systems, making it difficult to detect short-term anomalies that have an acute impact on public health. To address this issue, we modeled long-term trends in dengue incidence and quantified annual aberrations from those trends using a metric that we called RISc. This metric of dengue intensity quantifies the difference between reported incidence and expected incidence after removing global, regional, and country-level time-dependent trends from the data. As a result, we can use RISc to understand how the burden in 1990 compares to that of 2023 while accounting for differences in testing and case ascertainment.

RISc allows us to measure year-to-year fluctuations in dengue intensity, which is complementary to past efforts to quantify dengue burden, while also providing new opportunities to understand changing epidemiologic patterns. Many previous studies describe how dengue prevalence or incidence varies geographically by assigning static measures of transmission to identify regions with long-term dengue burden.^13,14,37^ However, with RISc we can detect year-to-year intensities independently of the long-term trend. This allows us to uniformly measure dengue burden relative to time- and location-specific contexts. To quickly identify anomalous years, we created three different intensity categories based on RISc, with instances in the top tier representing outbreak-like conditions and instances in the bottom tier representing abnormally low incidence years. In doing so, we have identified high and low intensity years in countries throughout the world using a uniform methodology. This is helpful because it allows us to conduct secondary analyses on global dengue patterns, despite differences in baseline transmission or case ascertainment geographically or over time.

We used RISc to understand country-level drivers of annual dengue intensity on global and regional scales through a meta-regression approach combining RISc values and several external mediating variables. Our analysis suggests that annual temperature anomalies impact intensity, with higher than usual temperatures associated with higher RISc (**Figure 5**). Local temperature is a powerful predictor of dengue incidence because it can directly modulate survival, development, biting rate, and dissemination rate amongst *Ae. aegypti* mosquitoes.^38,39^ Our finding that temperature anomalies, and ultimately global climate anomalies, drive short-term dengue incidence intensity is consistent with other studies.^7–9,36,38^ Furthermore, our finding that precipitation anomaly has no association with RISc is also consistent with other research; however, the effect of precipitation may be more pronounced at finer spatial scales.^36,38^ The impacts of precipitation on *Ae. aegypti* are complex and excessive precipitation can wash away breeding habitats leading to decreased transmission, while human water storage practices during droughts allow for abundant breeding sites and increased transmission.^38,40–^43

Additionally, we examined global and regional patterns of intensity using RISc. We found that recent years with high dengue burden may not have been unprecedented in terms of RISc. For example, while global case counts were much higher in 2023 than they were in 1998, the short-term anomalies of cases in relation to baselines (RISc) were similar between the two years (**Figures 1 and 4**). Epidemiologic reports from both years describe significant increases in incidence as well as concurrent outbreaks in many countries.^26,44–47^ Notably, global RISc was the highest in 1995, and this trend was largely driven by the Caribbean and Central America. Prior to 1995, many countries in these regions reported very few, if any, cases annually. Therefore, what is being captured is early emergence of dengue in these regions, which leads to an elevated RISc as the baseline used to determine RISc was very low.^26,48–50^ We also found that dengue intensity follows multi-year cycles, with groups of two or more years with positive RISc values grouping together, followed by groups of two or more years with negative RISc values (**Figures 3 and 4**). This observation is supported by our meta-regression analysis, which shows that if the previous year had a positive RISc, the next year was more likely to have a positive RISc (**Figure 5**). Taken together, this suggests that large-scale transmission dynamics are not as simple as having a one year outbreak that immediately leads to low-level transmission the year after. Rather, what we see is that high intensity years tend to fluctuate between multi-year cycles of positive and negative RISc. A possible explanation for this is population immunity and expected transmission dynamics.^46^ Dengue virus may circulate for several years, slowly building up population immunity until a critical threshold is reached at which conditions are no longer optimal for transmission.

Despite the insights we derived from this research, limitations remain. First, while we de-trended our data, we have not removed all forms of bias. This is particularly true in relation to data collection. We are missing data from many dengue endemic locations, including many Asian and African nations. This is largely a result of our inclusion criteria, which were set to favor locations with annual dengue recurrence throughout the time series. Therefore, we are not capturing all locations where dengue virus has recently emerged or where surveillance data has recently become available, such as Ethiopia, Cote d’Ivoire, and Nepal. Additionally, sudden changes to surveillance practices, such as active case-finding at the timepoint of emergence or during an outbreak, could artificially inflate RISc, while systematic lack of testing may yield an artificially low RISc. This may partially account for the low global intensity seen during the 2020-2021 seasons when the COVID-19 pandemic was at its peak (**Figure 4**). Further, the patterns of dengue virus transmission are governed by local factors, and because we are analyzing aggregated annual and national case data, we are likely missing some of the true subnational dynamics.^51,52^ This will have a disproportionate impact on larger dengue endemic countries and countries with diverse geographies like Brazil and Vietnam.^36,53,54^ Additionally, while we created intensity categories to quickly identify anomalous years and years with outbreak-like conditions, it is important to note that these categorizations are not based on outbreak definitions that are routinely used in public health applications. Dengue outbreak definitions are often set on district-wide or state-wide scales based on weekly or monthly case data with the explicit goal of triggering appropriate public health response, such as initiating vector control programs.^5^ Even among outbreaks described at national-levels in literature, there is significant fluctuation in regard to what degree of elevated incidence is needed to be considered an “outbreak” (**Figure S7b**).^2^ As a result of using annual national-level data, our use of RISc presented here is a summary measure and does not infer within-season or subnational dynamics. Further, it is important to note that the leading edge of RISc estimates and intensity classifications may change with the inclusion of future data, as they would further inform the baseline trend from which RISc is derived. Finally, data on covariates used in the meta-regression analysis were aggregated to national and annual scales, which could obscure important correlations with intensity at finer geographic and temporal resolutions.

In sum, our study provides a retrospective analysis of global, regional, and country-level dengue intensity dynamics and drivers over the past 30 years. Our raw data, as well as the results of our RISc analysis, are open-source and can be used as starting points for future research to further explore dengue epidemiology or evaluate forecasting models. Future research could look to apply this framework on finer scales, as separating short-term transmission anomalies from long-term trends would be particularly useful in understanding how local impacts of climate and population immunity modulate year-to-year transmission. It could also be used in public health decision-making to identify short-term excess burden and allocate resources appropriately, or to better characterize long-term burden and plan strategic multi-year risk reduction programs. Finally, the challenges addressed in this study are not unique to dengue, and therefore, the approaches we present here could be applied to other infectious disease systems.

## Methods

### Data acquisition and refinement

To analyze global, regional, and national dengue trends, we compiled a dataset with surveillance, population, and geographic data for countries with the highest dengue burden. We assessed surveillance data from OpenDengue, WHO, PAHO, Gideon, and Asian Dengue Voice and Action (ADVA) for completeness and consistency between datasets.^1,2,22,23,55^ OpenDengue, WHO, and PAHO had the greatest alignment of reported cases across the individual datasets. Gideon data, which is largely derived from scientific literature, often had the finest geographic resolution but had the most gaps in data and was the least consistent with other data sources. Similarly, ADVA data had many gaps not seen in OpenDengue, WHO, and PHO datasets. OpenDengue had more uniform reporting in Asia than WHO and was therefore the most complete single source of information and was used to obtain case data for all analyses. National population data were obtained from The World Bank, joined to OpenDengue data, and used to calculate incidence rates.^25^ Finally geographic information denoting the region/continent of each country was obtained from R Natural Earth.

From the OpenDengue dataset, we selected national-level data on cases from 1990-2024, which was reported either annually or sub-annually depending on availability. When weekly or monthly data were available in the southern hemisphere, the dengue season was offset 6 months from the calendar year to start in July and end in June to account for the peak transmission that often occurs between December-January in this region. For countries that lie on the equator, the centroid of the country was used to determine the hemisphere. As a result, our final dataset includes dengue seasons from 1990-2023. We then subset the data to include any location that had reported 0 cases or had missing data for 15 or fewer years. Any location that was missing population data through the World Bank was excluded. We further selected for countries that had either 1) an average of 500 or more cases annually over the time series; or 2) an incidence rate of 10 or more cases/100,000 population annually over the same time frame. This resulted in a dataset that had annual dengue incidence for 57 countries over 34 years with 1,995 observations of dengue case data and 43 country-year combinations with no reported data (**Figures 1 and S1**).

Data on external covariates used in the meta-regression analysis were obtained from the World Bank and Copernicus Climate Change Service (C3S). Specifically, the World Bank World Development Indicators dataset was used to obtain information on GDP per capita (indicator code (IC): NY.GDP.PCAP.PP.KD), forest area (IC: AG.LND.FRST.ZS), access to electricity (IC: EG.ELC.ACCS.ZS), and urbanicity (IC: SP.URB.TOTL.IN.ZS).^25^ Hourly temperature and precipitation data aggregated by month were obtained from the C3S ERA5 reanalysis dataset.^56^ The geocoded climate data was fit to country boundaries obtained from R Natural Earth and the monthly mean across each country’s polygon was extracted.^57^ This data was then aggregated to the annual level, matching the dengue season offset in the southern hemisphere, when applicable. Finally, the annual average from 1991-2020 for each country was calculated and used as the reference value to calculate the temperature and precipitation anomalies.

### Developing RISc values and extracting long-term trends

We analyzed the dengue case data using a hierarchical Bayesian Poisson regression model (**Figures 2, S2-6, and S8-9)**). The model was fitted in R using the Integrated-Nested Laplace Approximation framework for inference using the R INLA package.^58^ We denote *Y*_*i*,*t*_ as the observed number of cases in country *i* and year *t*, and *E*_*i*,*t*_ as the population (offset). We assumed *Y*_*i*,*t*_ to be distributed as *Poisson*(*E*_*i*,*t*_*λ*_*i*,*t*_). We then model the log-incidence as:

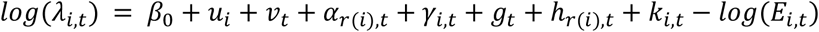

Here, *β*_0_ is an intercept; u_i_ and v_t_ are country- and year-specific random effects (iid Gaussian); *⍺*_r(i),t_ and *γ*_i,t_ capture interactions as unstructured region-year and country-year random effects; and g_t_, h_r(i),t_, and k_i,t_ are smooth temporal trends at the global, regional, and country levels, each modeled using a second-order random walk (RW2).^59^ Overdispersion in the surveillance data was accounted for through the inclusion of the random effects above. This structure enabled the model to flexibly account for abrupt aberrations (*u*_*i*_, *v*_*t*_, *⍺*_*r*(*i*),*t*_, *γ*_*i*,*t*_), gradual changes over time (*g*_*t*_, *h*_*r*(*i*),*t*_, *k*_*i*,*t*_), and regional differences (*⍺*_*r*(*i*),*t*_, *h*_*r*(*i*),*t*_), while the offset (*E*_*i*,*t*_) adjusted for underlying population size. All components were estimated jointly in a Bayesian framework using Integrated Nested Laplace Approximation (INLA), which is an alternative to traditional MCMC methods. The priors for the iid terms were the uninformative Gaussian priors (INLA default) while the priors for the random-walk terms were the penalized complexity priors defined as Gaussian distribution with weak precision (also INLA default). Our model was validated using a range of informative and uninformative priors, none of which led to substantial changes in RISc or conclusions from the meta-regression analysis (**Figures S11 and S12**).

Imputed values were validated by removing known data that fit the general pattern of missing data, re-running the model, and examining the Pearson correlation coefficient between the known and imputed values (**Figure S8**). When data were absent, we mapped the pattern of missingness by identifying (1) the average number of countries per region with missing data; (2) the average number of years with missing data per country; and (3) the average number of consecutive years of missing data. We then randomly selected data matching this pattern from regions without missing data (Central and South America) to complete the validation as described.

Country-specific RISc values were then calculated by summing the mean estimates of the time-varying random effects, terms g_t_, h_r(i),t_, and k_i,t_. Intensity categories were created by balancing the specificity and sensitivity of high and low intensity years (**Figure 3**). With the RISc values centering on 0, we set values greater than or equal to 0.5 as high intensity (roughly 65% increase in cases above baseline), values less than or equal to -0.5 as low intensity (roughly 40% decrease in cases below baseline), and any value in between as expected intensity. This means a high RISc indicates years where we then compared this calibration against outbreaks declared by the WHO and reported in literature to assess how well this system captured outbreaks described in practice (**Figure S7**).^2,26^ All six dengue outbreaks declared by the WHO since 2005 in countries included in this analysis were used in the comparison. To obtain data on outbreaks described in the literature, we randomly selected 3-4 countries per region and then identified national-level outbreaks summarized in the Gideon dengue-specific literature review.^2^ Global and regional RISc values were calculated as the average RISc per dengue season across all countries (**Figure 4**). Long-term trends in incidence at the country-level were derived by multiplying the fitted values from the model by the country-specific population and dividing by the exponentiated RISc value, thus removing the random effects from the fitted values to show the remaining long-term trends estimated by the second-order random walk terms included in the model.

### Meta-regression analyses

Meta-regression analyses were performed using the R metafor package to identify large-scale environmental and socioeconomic associations with RISc (**Figure 5**).^60^ We used the posterior means and posterior standard deviations for RISc as the inputs to the meta-regression model. Correlation amongst covariates was assessed to ensure minimal collinearity (**Figure S10**). These covariates were scaled and put into the regression model along with RISc as the outcome and its associated variance, and were analyzed using a restricted maximum likelihood estimation method. This process was repeated once to analyze the data globally and once to analyze the data regionally by including region as an interaction term with the other candidate mediators. To extract region-specific estimates and confidence intervals, point estimates for each region were added to the point estimates from our reference region (Asia).

### Statistical analysis

All analyses were performed using R Statistical Software (v4.4.2).^61^ We used the vroom, tidyverse, lubridate, sf, stringr, janitor, rnaturalearth, exactextractr, metafor, and INLA R packages.^57,58,60,62–68^

## Data Availability

All data produced in the present study are available upon reasonable request to the authors

https://opendengue.org/

https://data.worldbank.org/

https://cds.climate.copernicus.eu/datasets/reanalysis-era5-single-levels?tab=overview

## Acknowledgements

We acknowledge Isabella Gamez for data curation, T. Alex Perkins for model advice, and S. Taylor, P. Jack, and Charly G. for technical discussions. This publication was made possible by the National Institute of Allergy and Infectious Diseases of the National Institutes of Health (NIH) under Award Number DP2AI176740 (NDG) and 5F31AI186435 (AJP) as well as pilot funds provided by Yale Planetary Solutions (via the Three Cairns Climate Impact Innovation Fund and the Office of the Provost AI Initiatives) and the Ambrose Monell Foundation (NDG and CJC). The findings and conclusions in this report are those of the author(s) and do not necessarily represent the official position of the NIH.

## Author Contributions

A.J.P., D.M.W., and N.D.G. designed the study. A.J.P., D.M.W., N.D.G., C.J.C., and J.L.W. developed the analytical framework. A.J.P. and R.L. analyzed the data. D.M.W., C.J.C., and J.L.W. provided statistical guidance. A.J.P. wrote the first draft of the manuscript. All authors interpreted and discussed the results. A.J.P., R.L., C.J.C., O.J.B, D.M.W., and N.D.G. discussed the epidemiological implications. All the authors edited and approved the contents of the manuscript.

## Supplementary Materials

### Supplementary Figures

**Figure S1.**
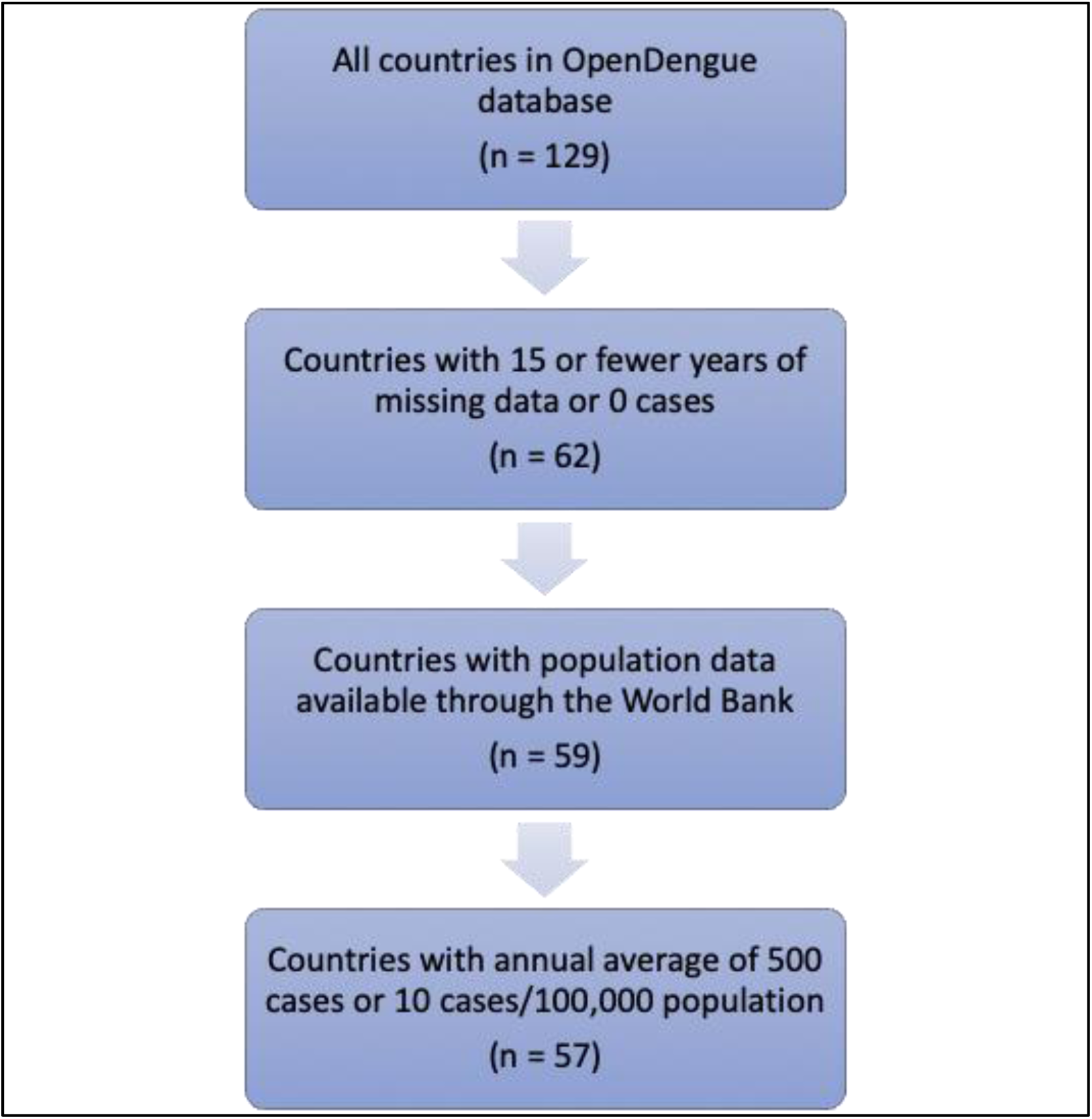
Selection of countries based on our inclusion/exclusion criteria. The majority of countries were excluded when selecting countries with a minimum of 19 years of data available between 1990-2023. Three countries were excluded because population data was not available through the World Bank and two countries were excluded for having too few annual cases.

**Figure S2.**
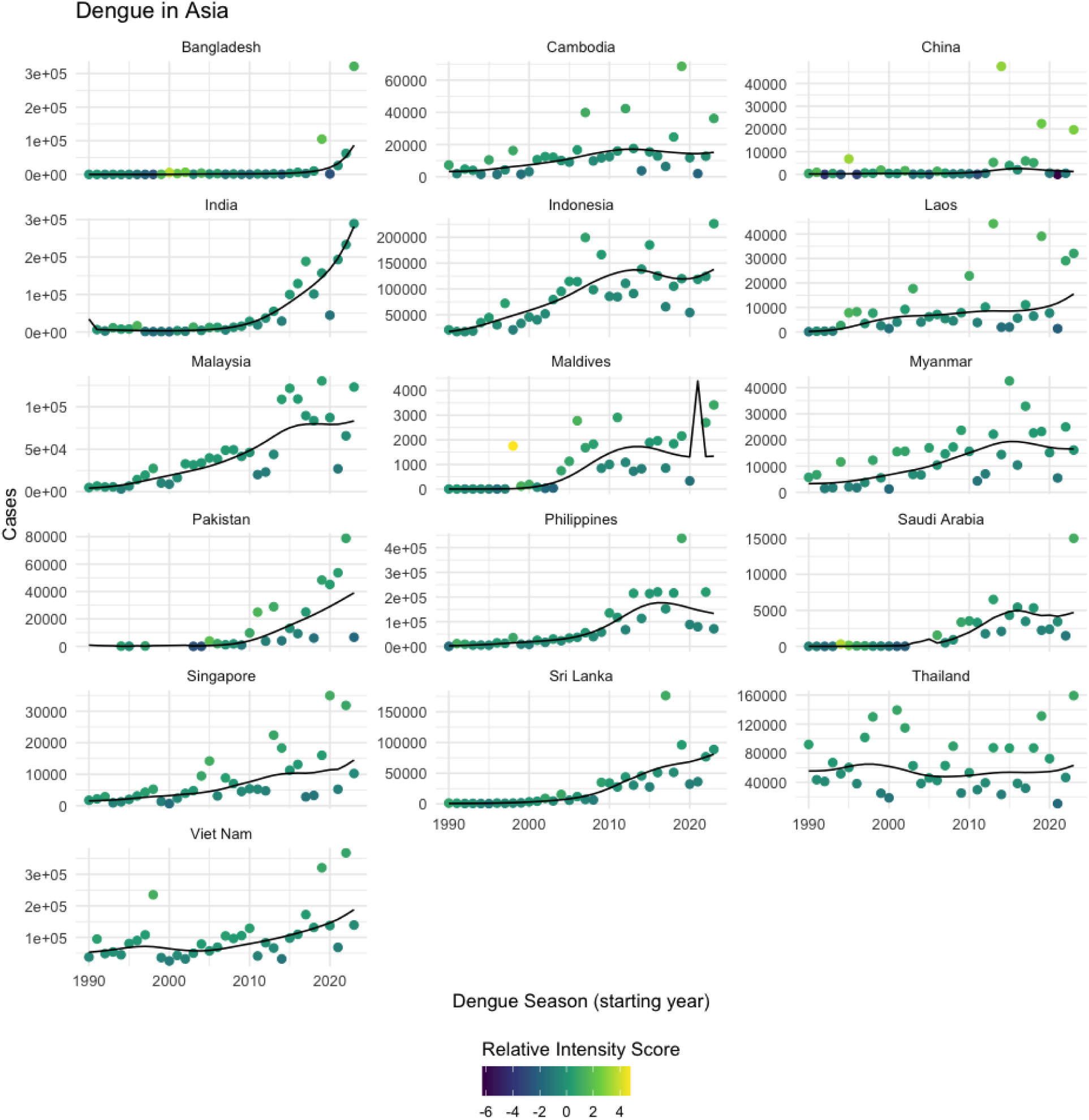
Annual RISc for all countries from Asia included in the analysis.

**Figure S3.**
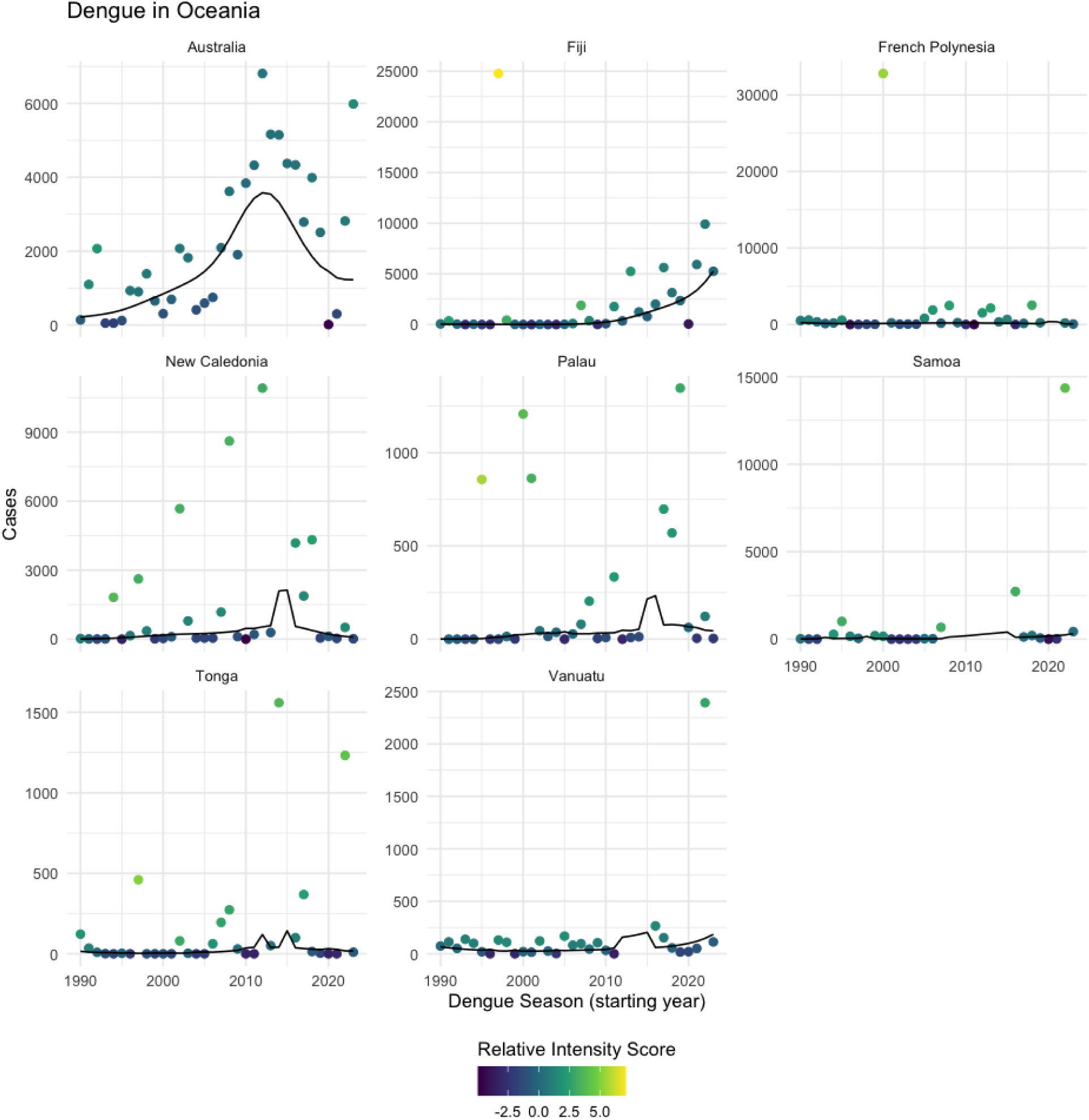
Annual RISc for all countries from Oceania included in the analysis.

**Figure S4.**
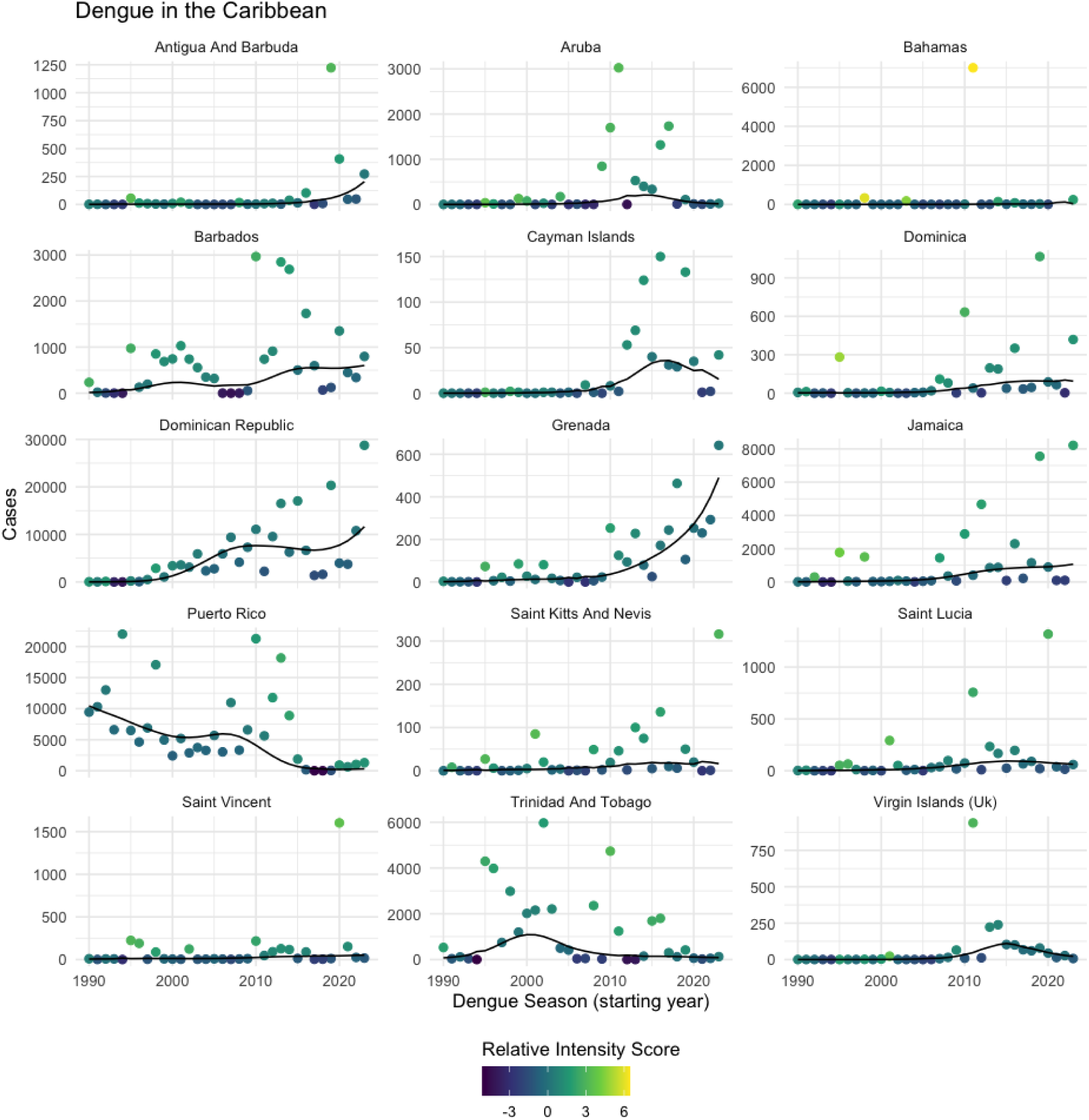
Annual RISc for all countries from the Caribbean included in the analysis.

**Figure S5.**
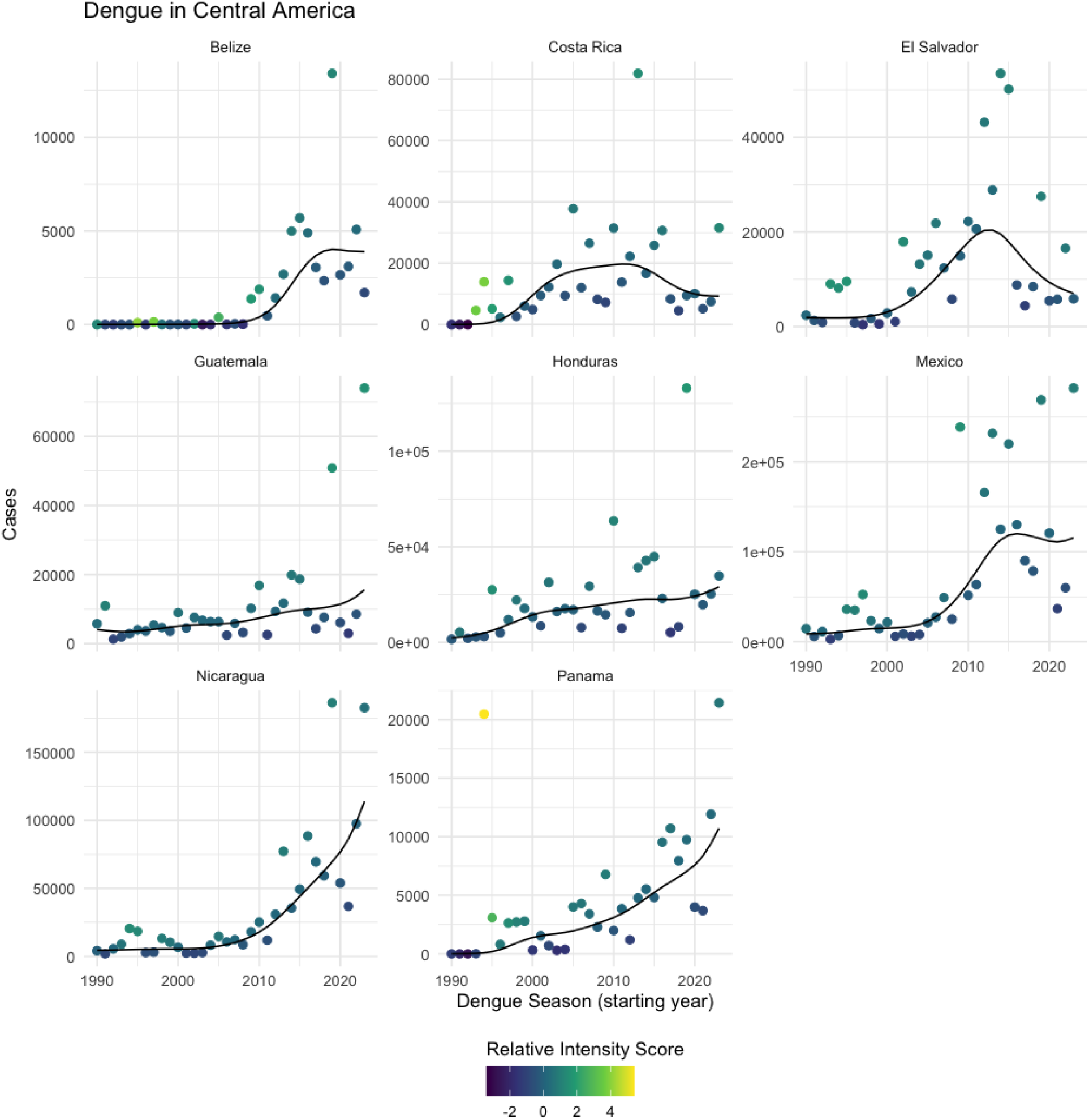
Annual RISc for all countries from Central America included in the analysis.

**Figure S6.**
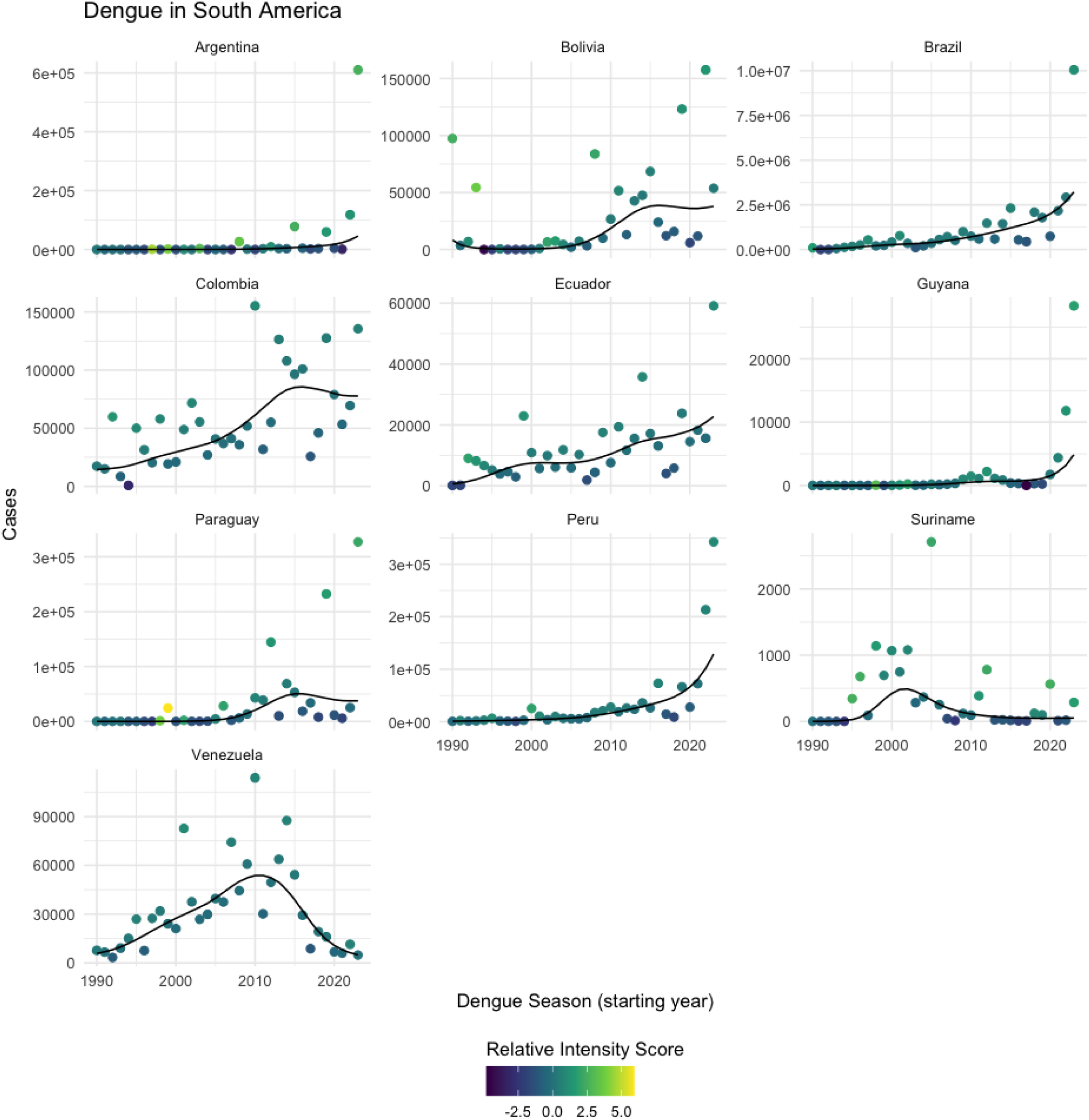
Annual RISc for all countries from South America included in the analysis.

**Figure S7.**
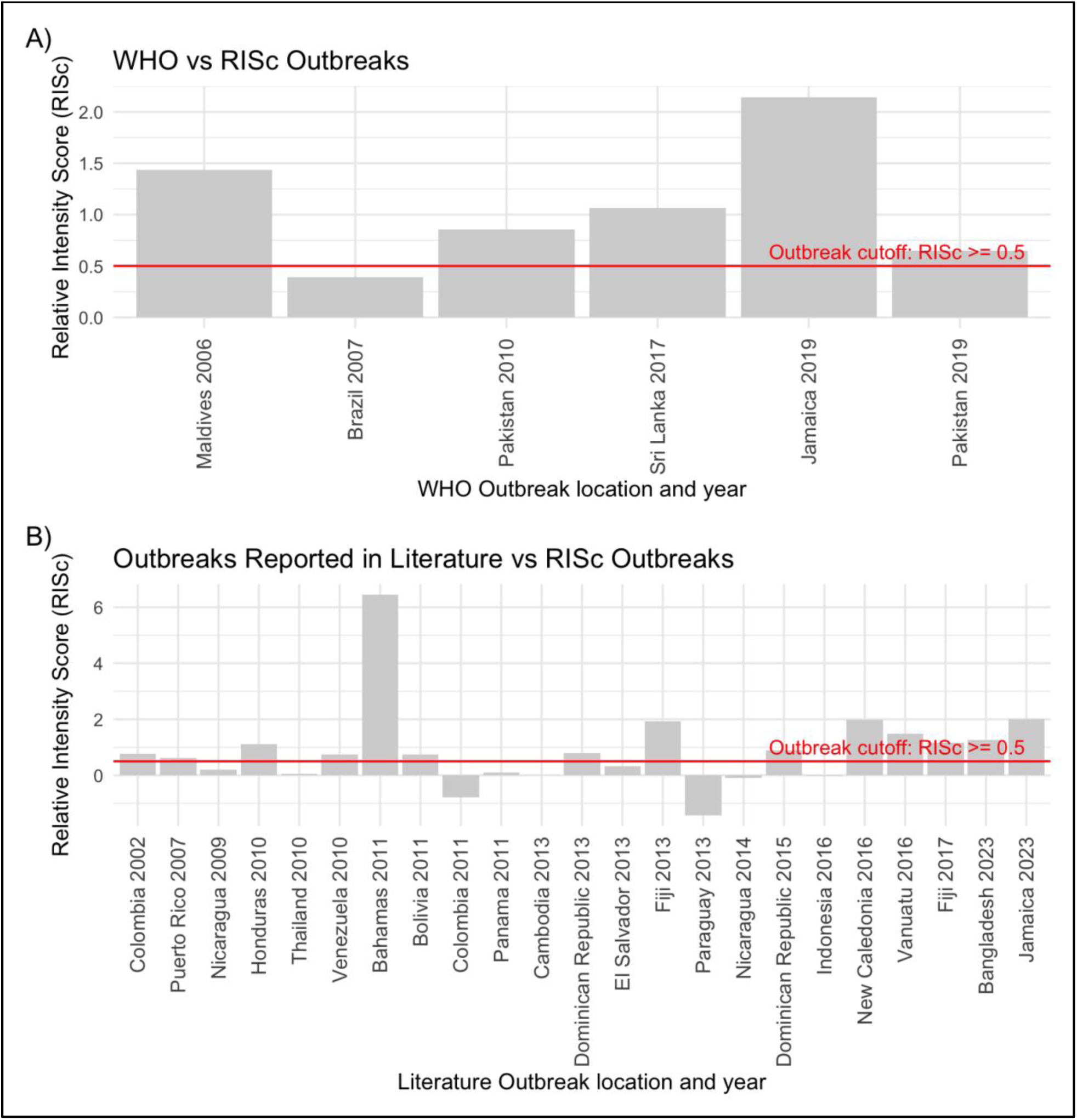
Comparison of high intensity RISc classification against declared dengue outbreaks. The red lines show the threshold value for “high intensity” transmission. (**A**) RISc of WHO declared dengue outbreaks between 2005-2022. (**B**) RISc of outbreaks declared in literature. Three to four countries from each region were randomly selected for inclusion in this comparison.

**Figure S8.**
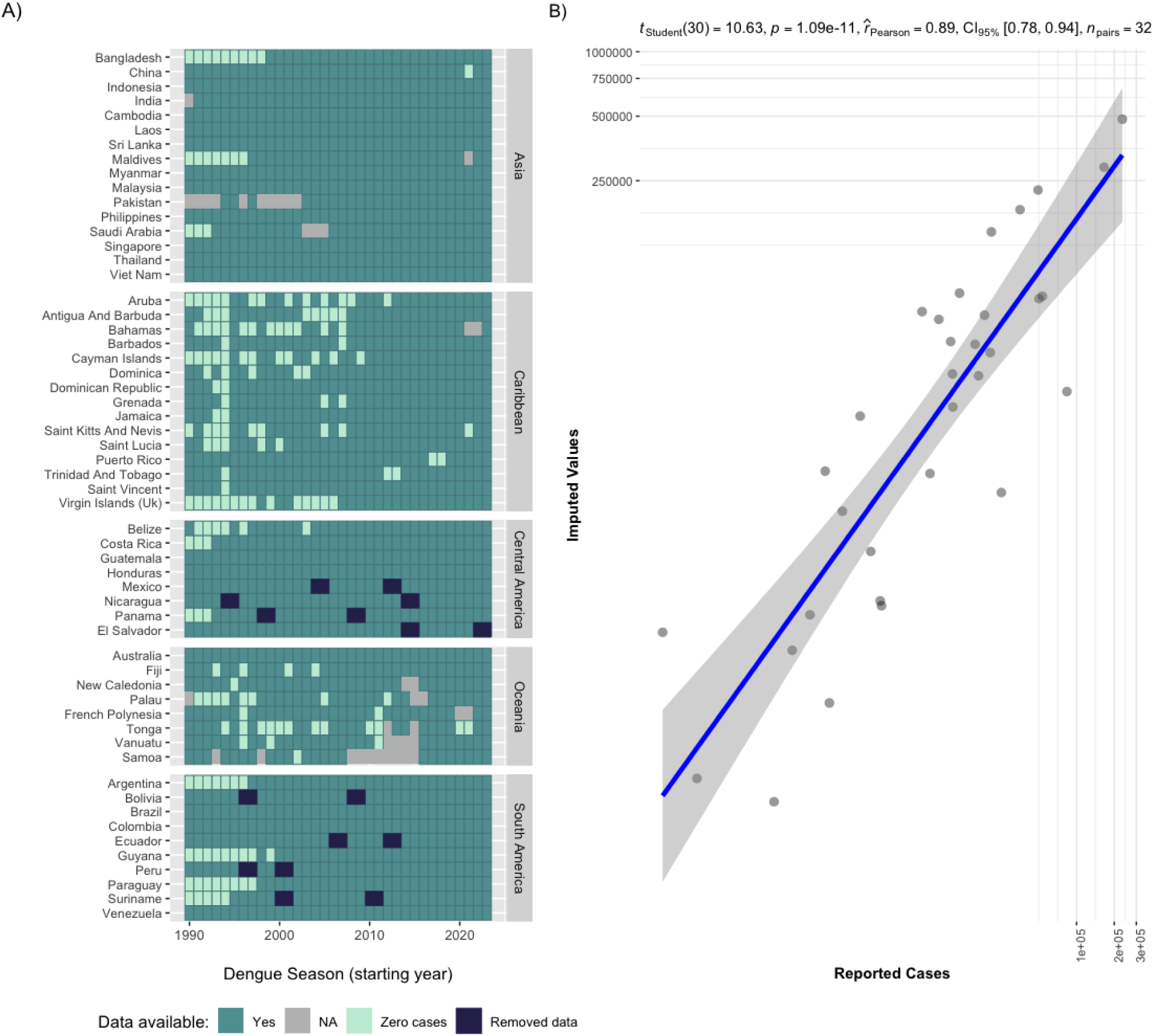
Validation of imputed values. A) The pattern of missing data (grey tiles) was identified in Asia, the Caribbean, and Oceania. Data fitting this pattern was then removed (dark blue tiles) from Central and South America. B) The correlation between known and imputed values is shown on the left.

**Figure S9.**
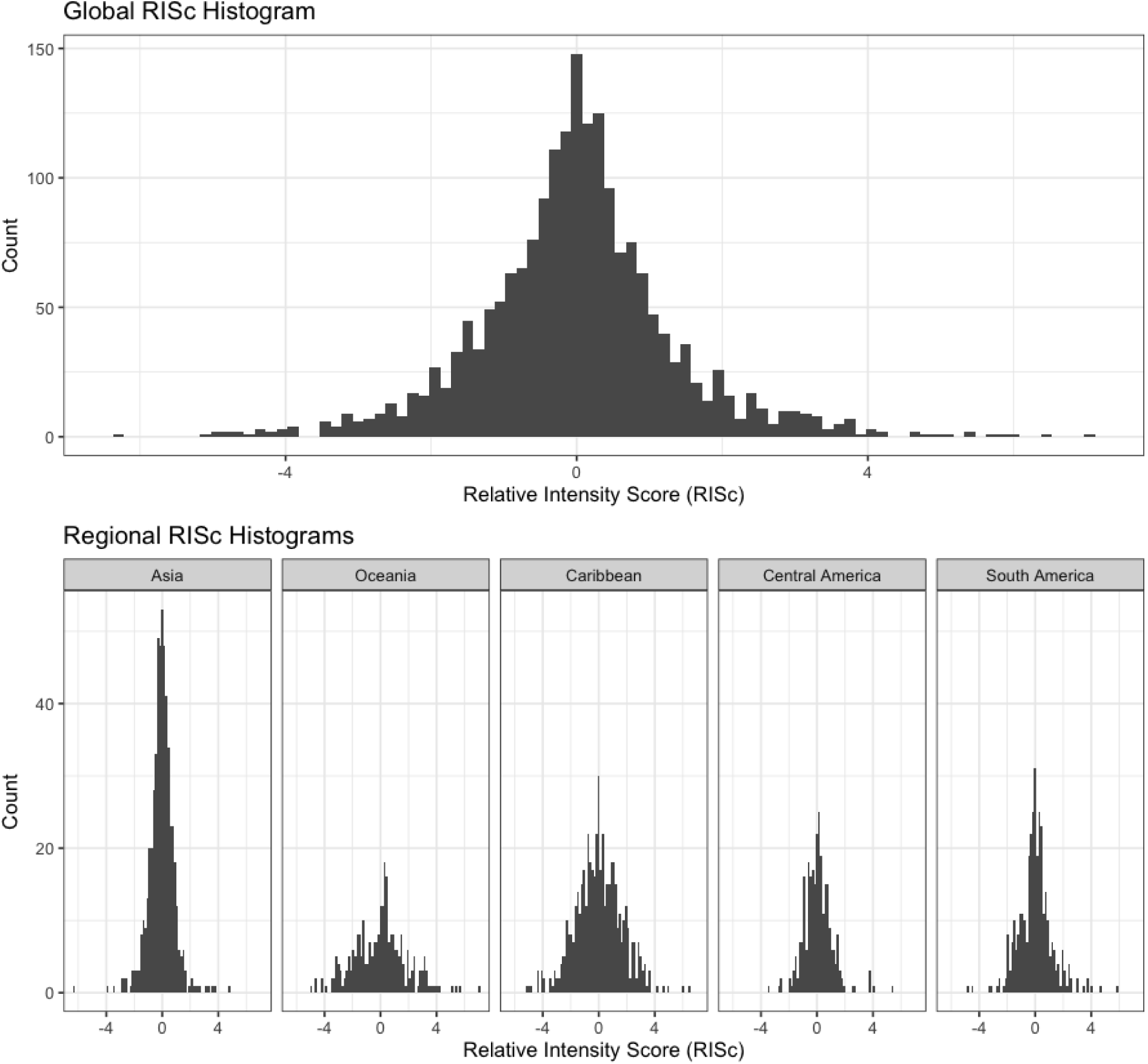
Histograms of RISc values globally and regionally.

**Figure S10.**
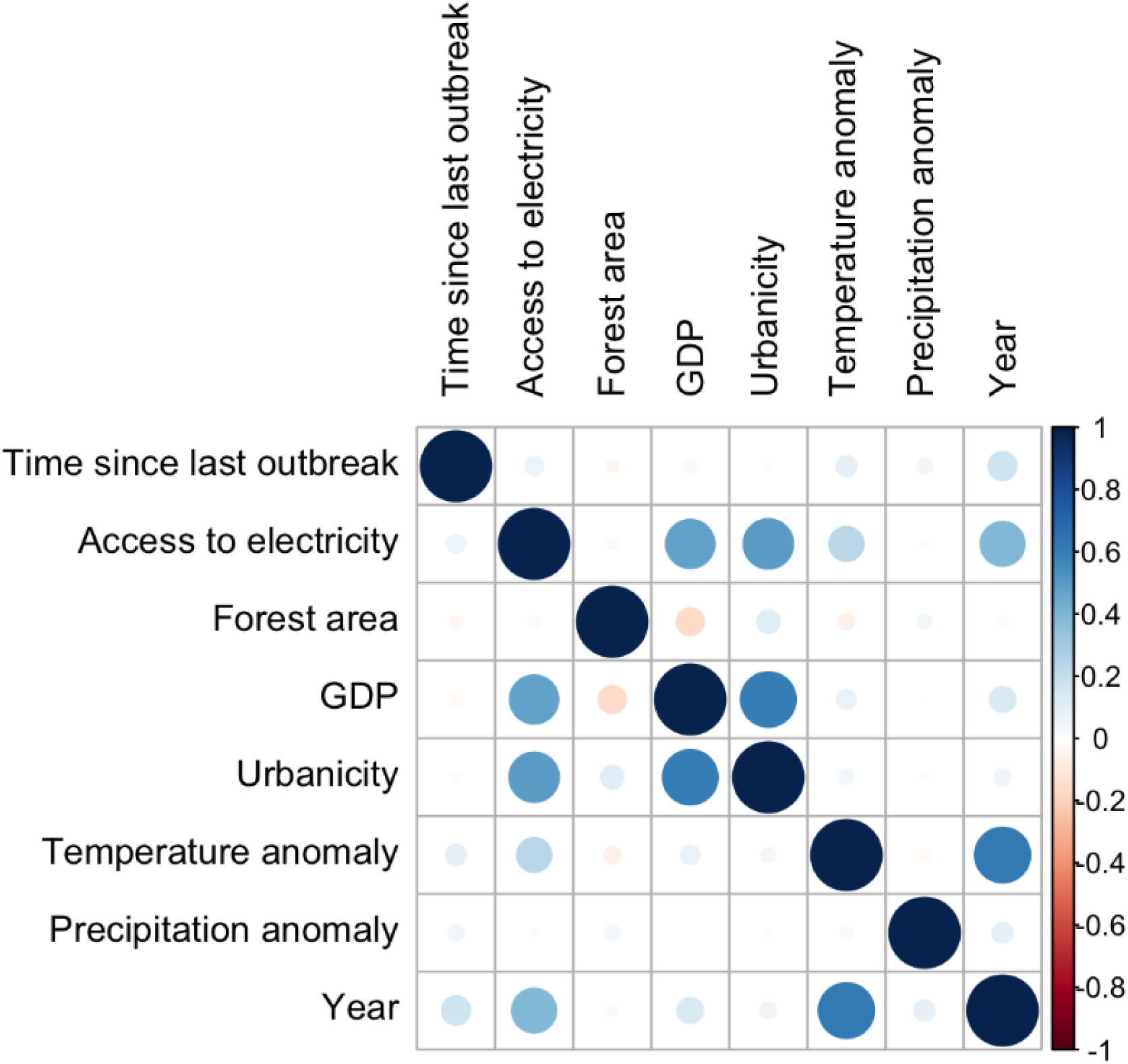
Correlation plot of covariates included in GLM analysis.

**Figure S11.**
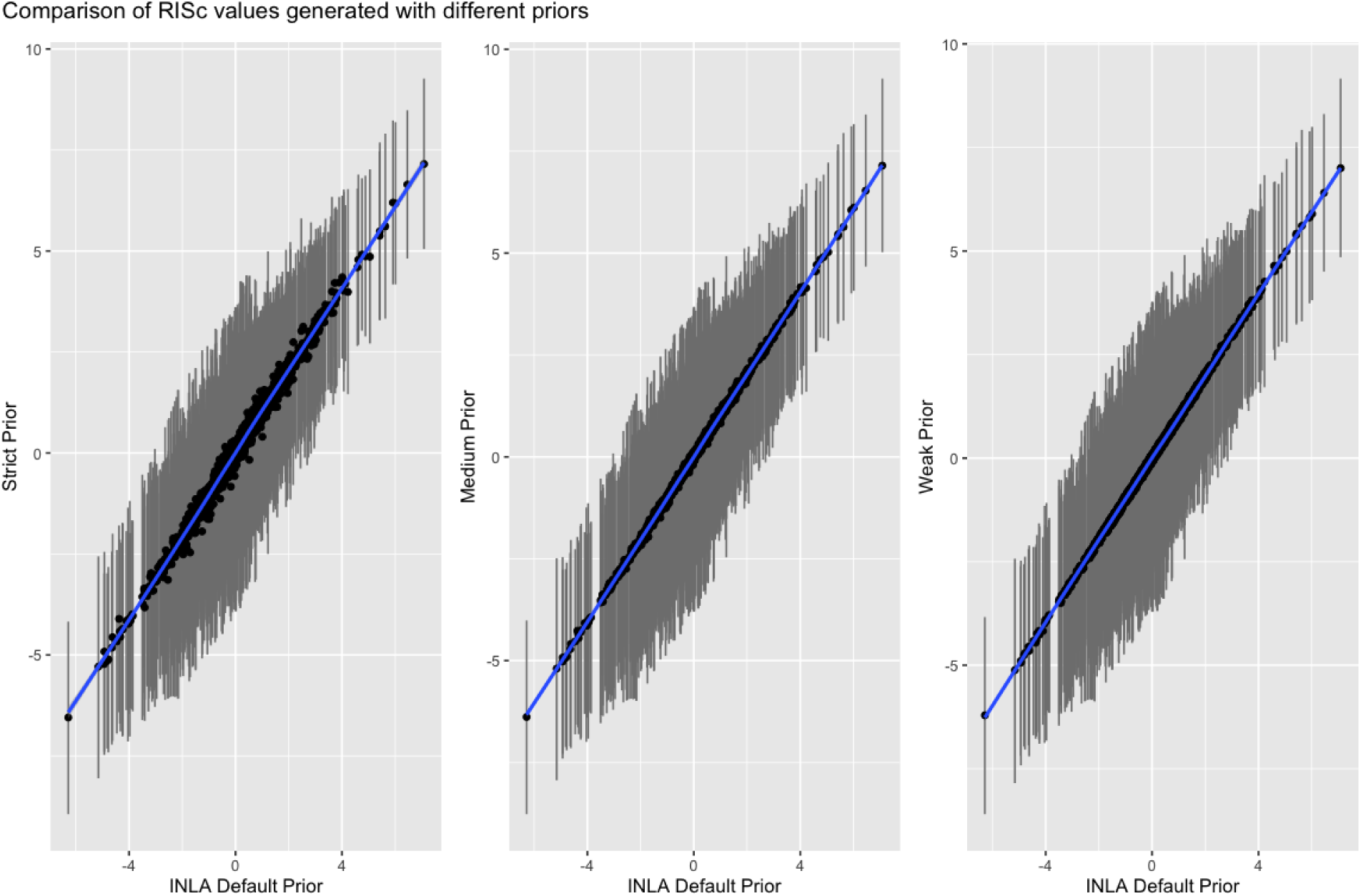
Comparison of penalized complexity priors. The correlation outputs between the INLA default priors as compared to a range of informative and uninformative priors used to inform the second order random walk components of our model. Point estimates for RISc are shown as black dots and the 95% confidence intervals for each point are shown as grey lines.

**Figure S12.**
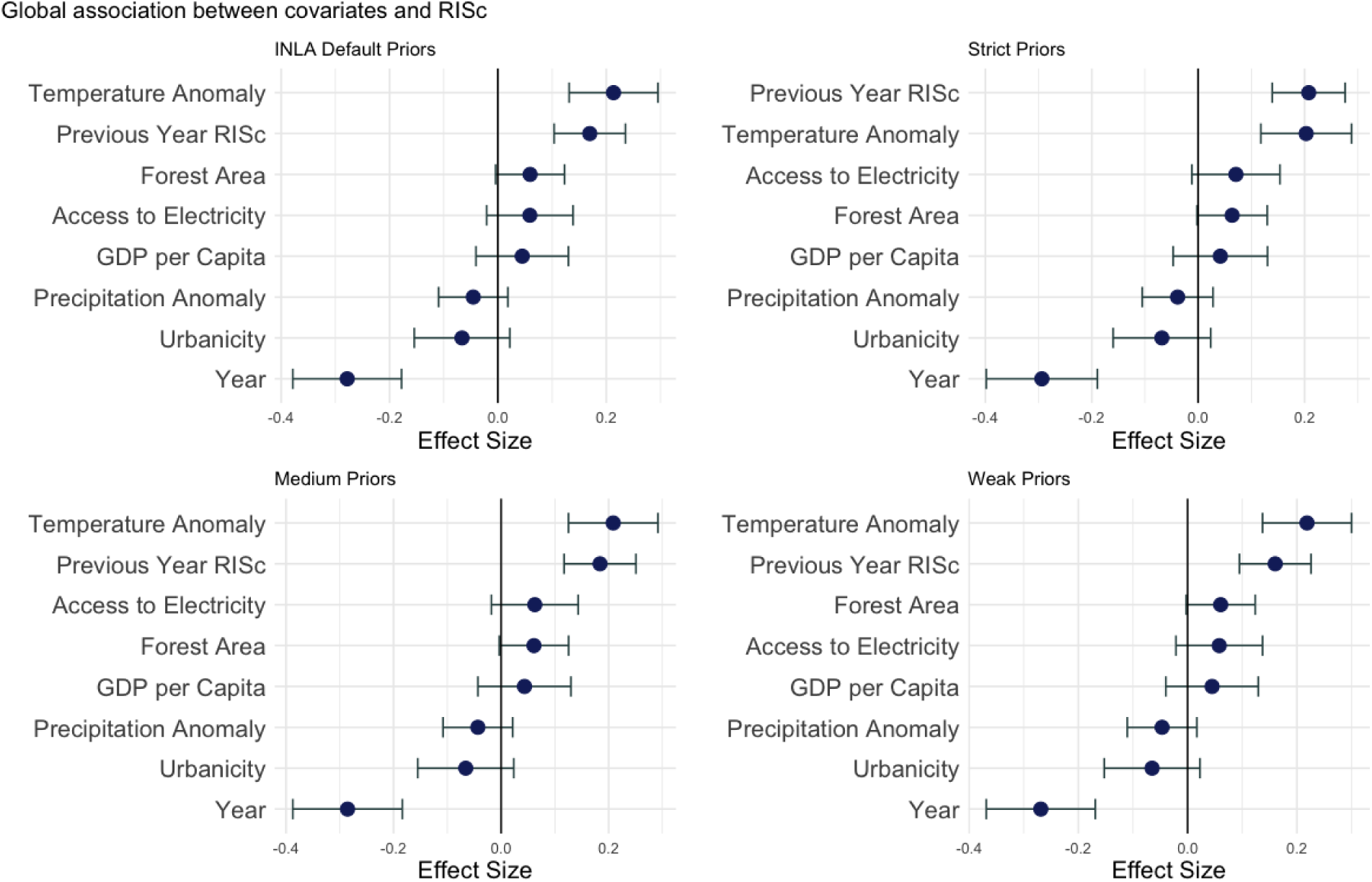
Effects of multiple penalized complexity priors on the meta-regression analysis. Changes to the priors used to inform smoothing terms in the Bayesian Poisson regression model lead to no substantial differences to the conclusions drawn from the meta-regression analysis.

