## Supplementary figures and images for "Standardizing annual dengue intensity reveals global drivers of transmission"

### Figure S1

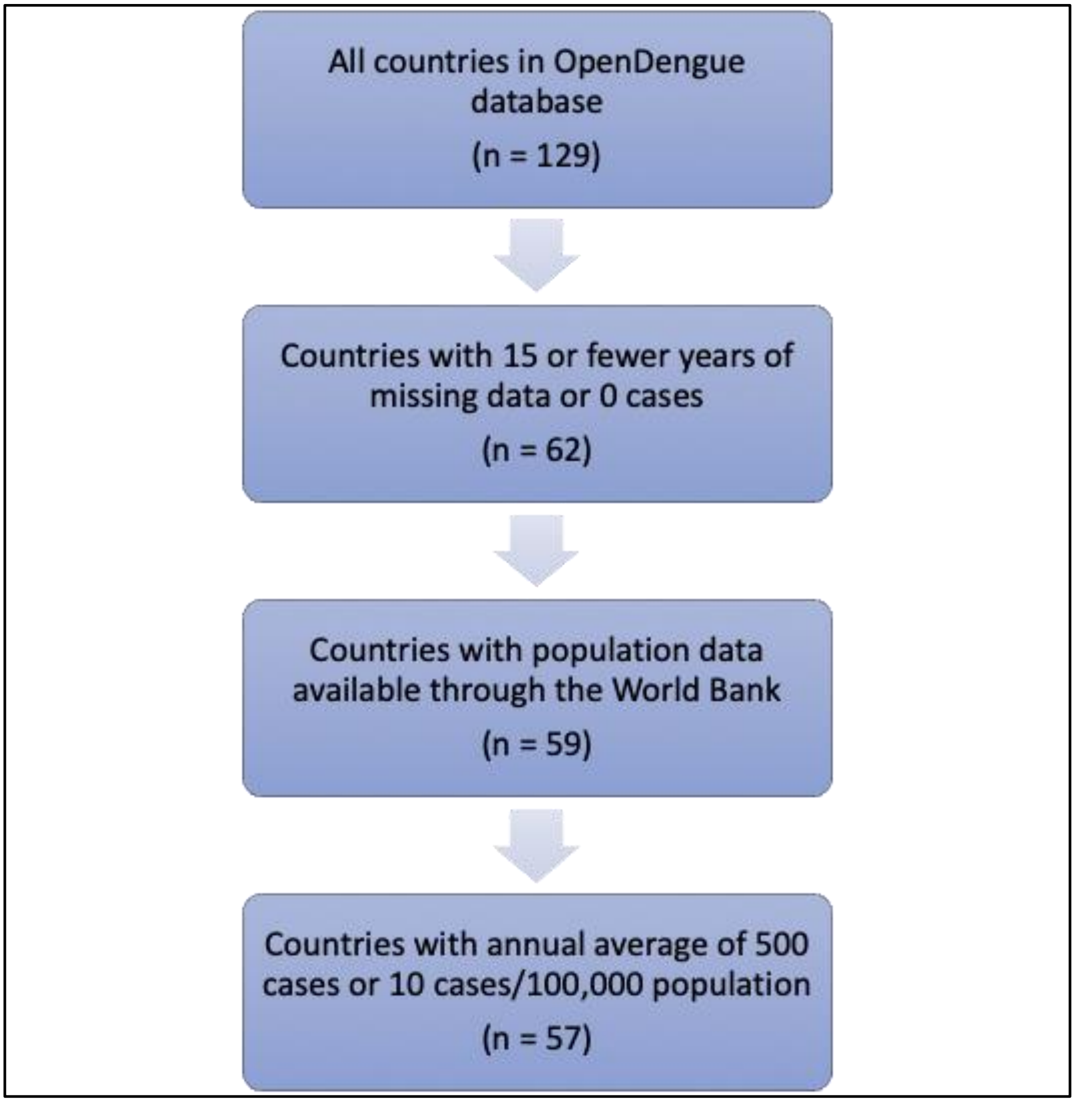

### Figure S2

#### Dengue in Asia

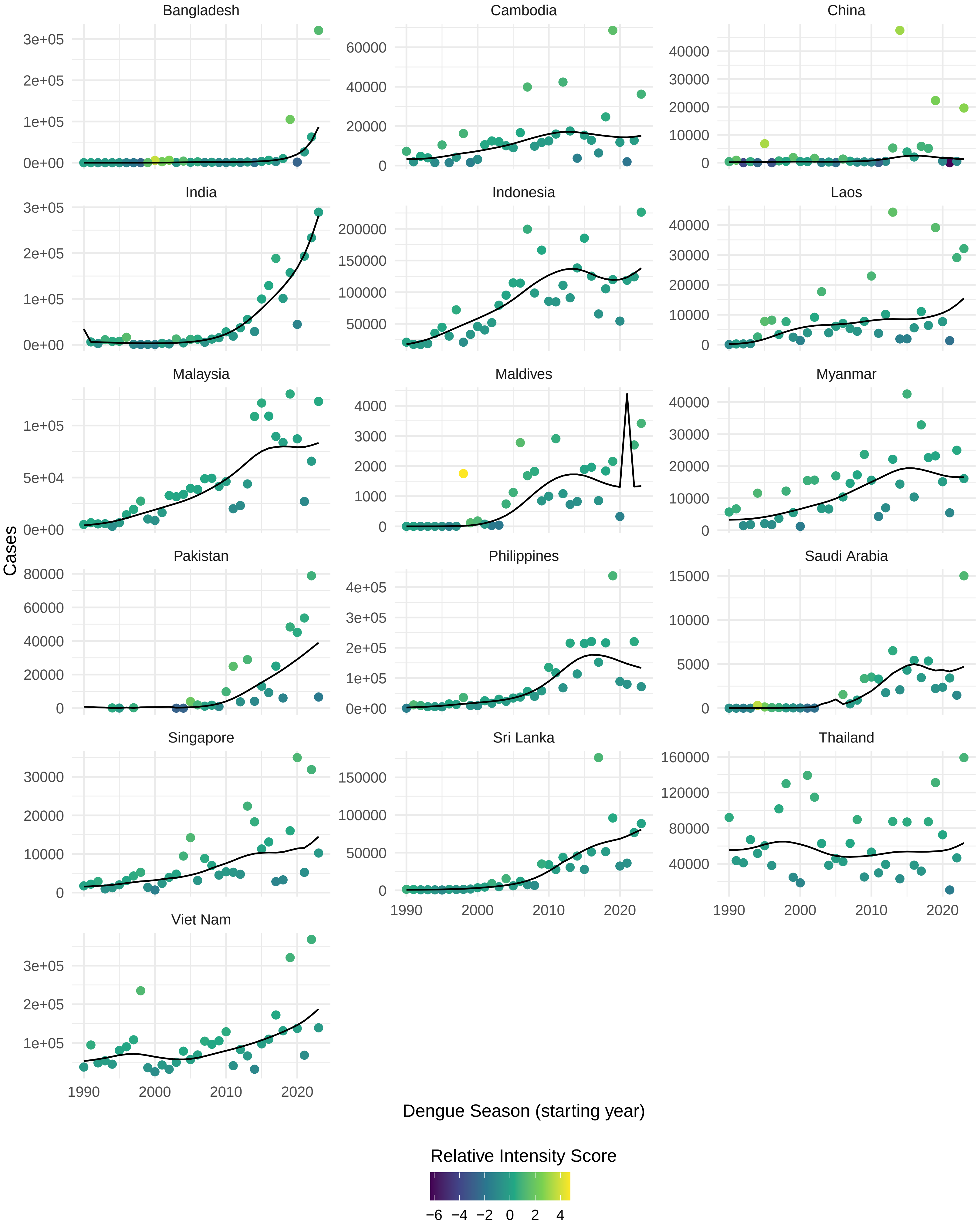

### Figure S3

#### Dengue in Oceania

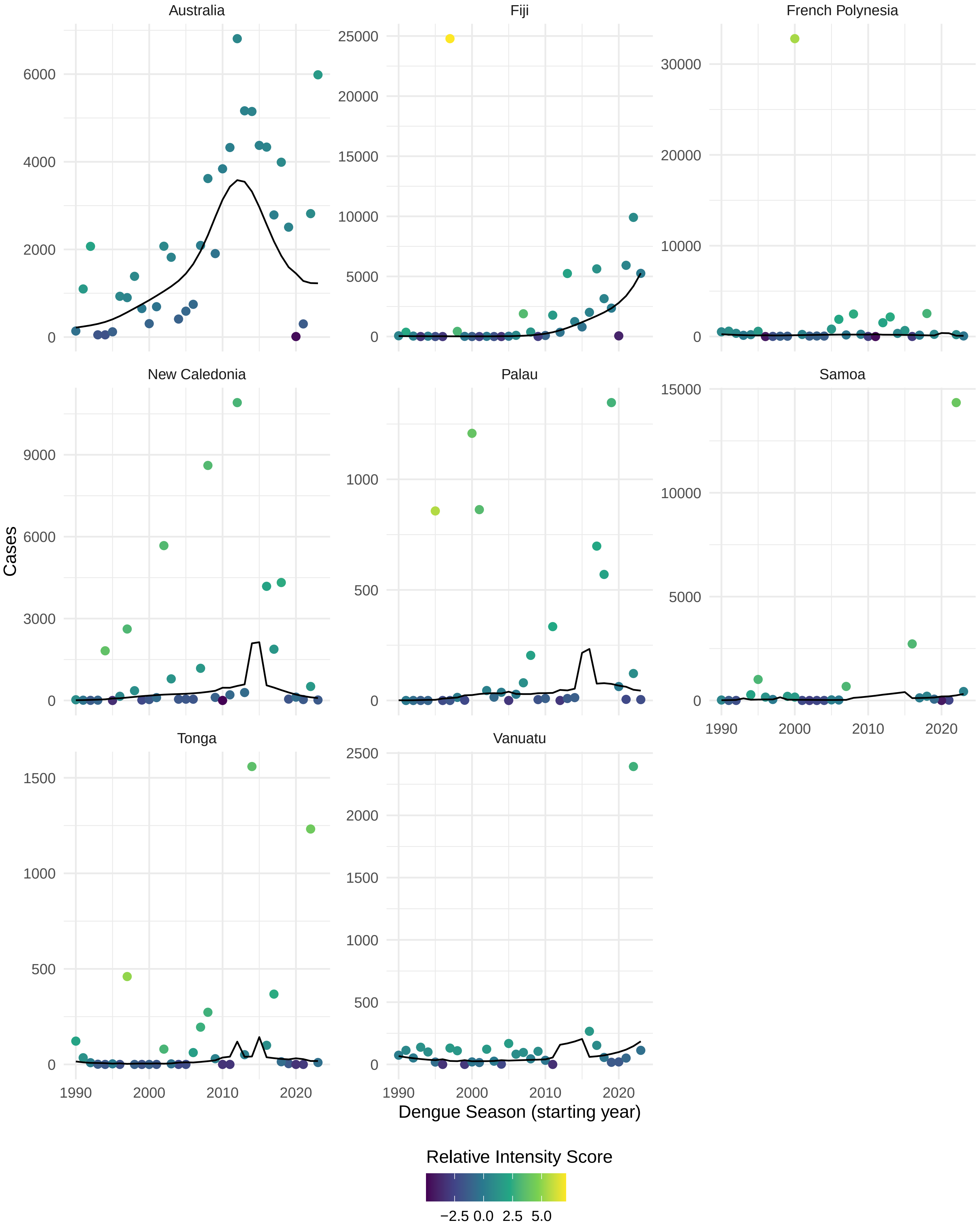

### Figure S4

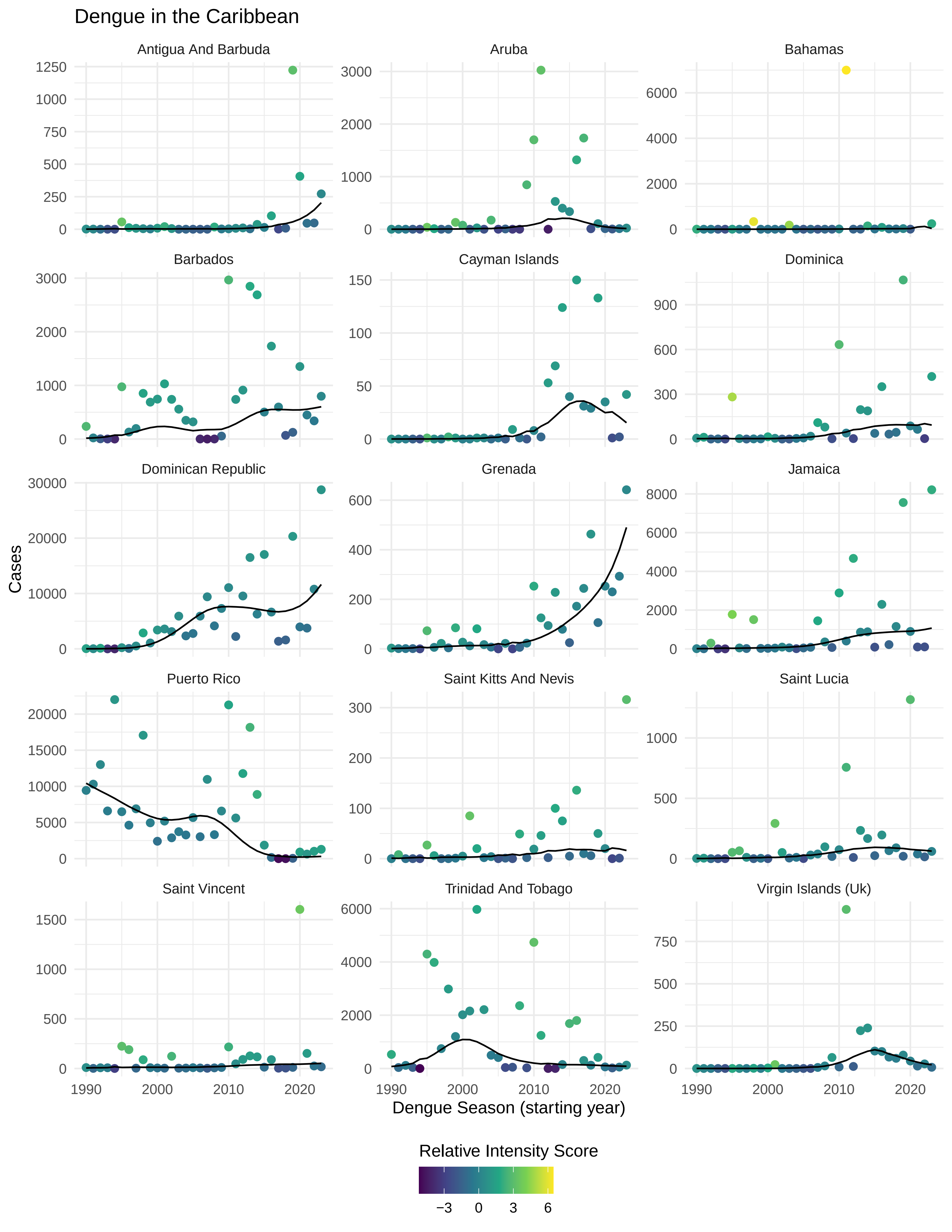

### Figure S5

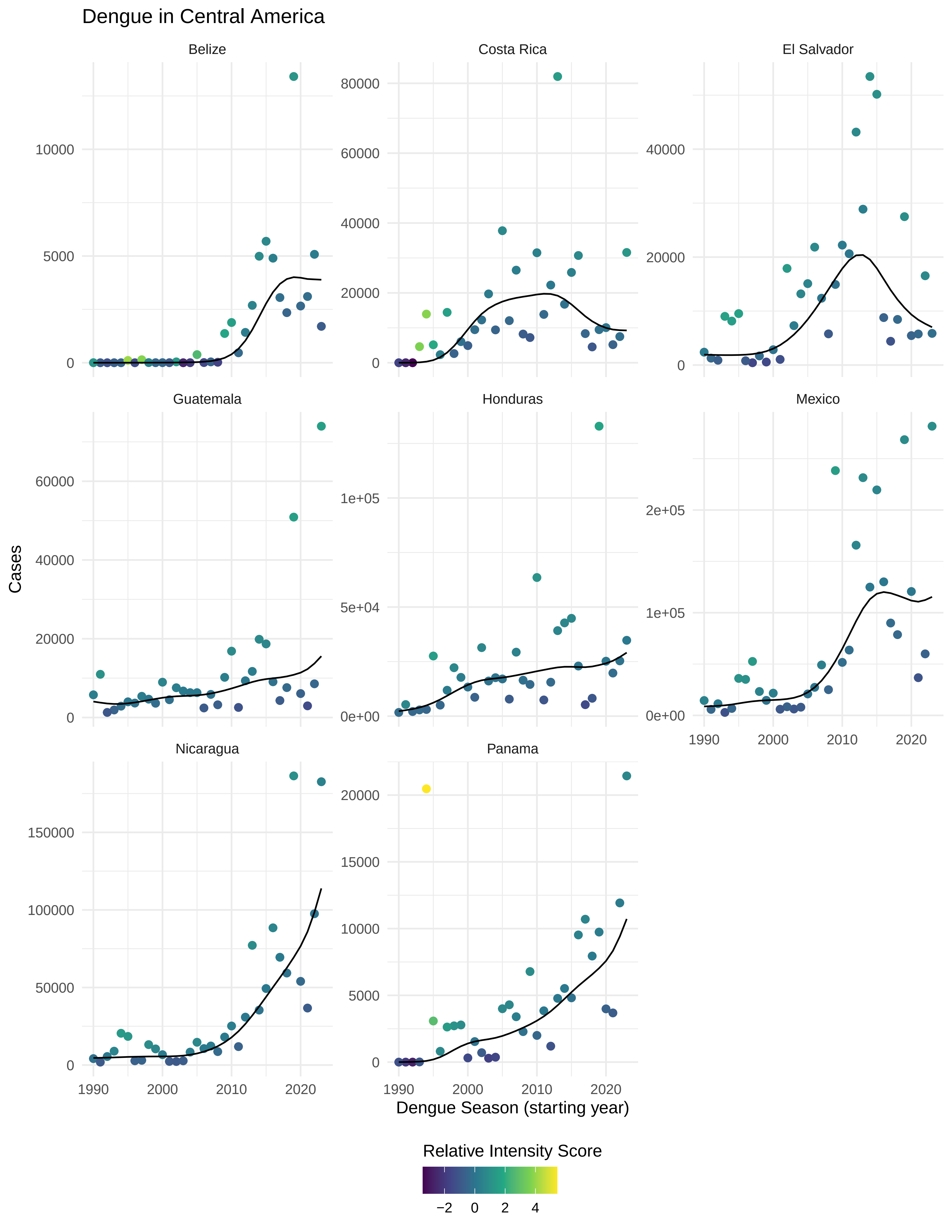

### Figure S6

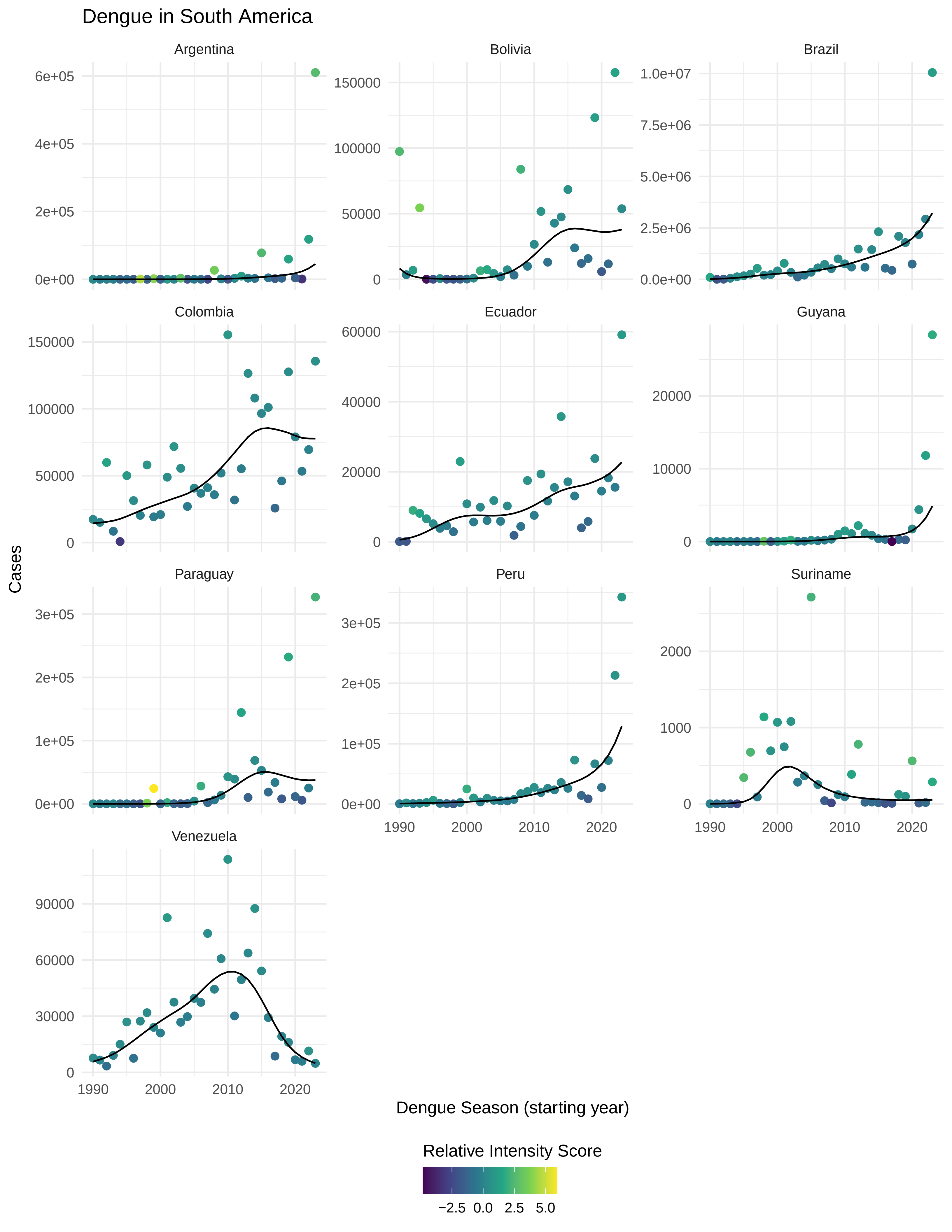

### Figure S7

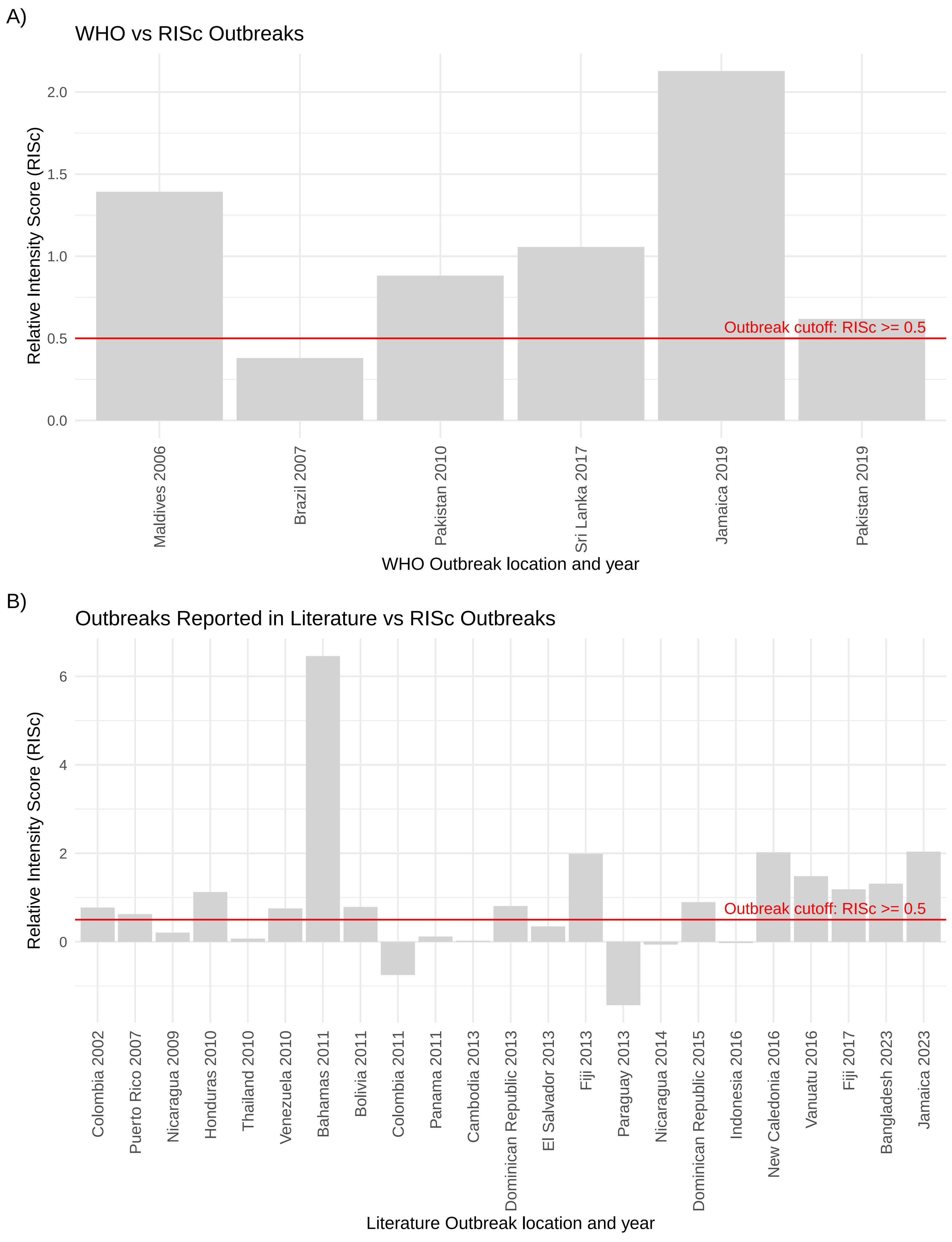

### Figure S8

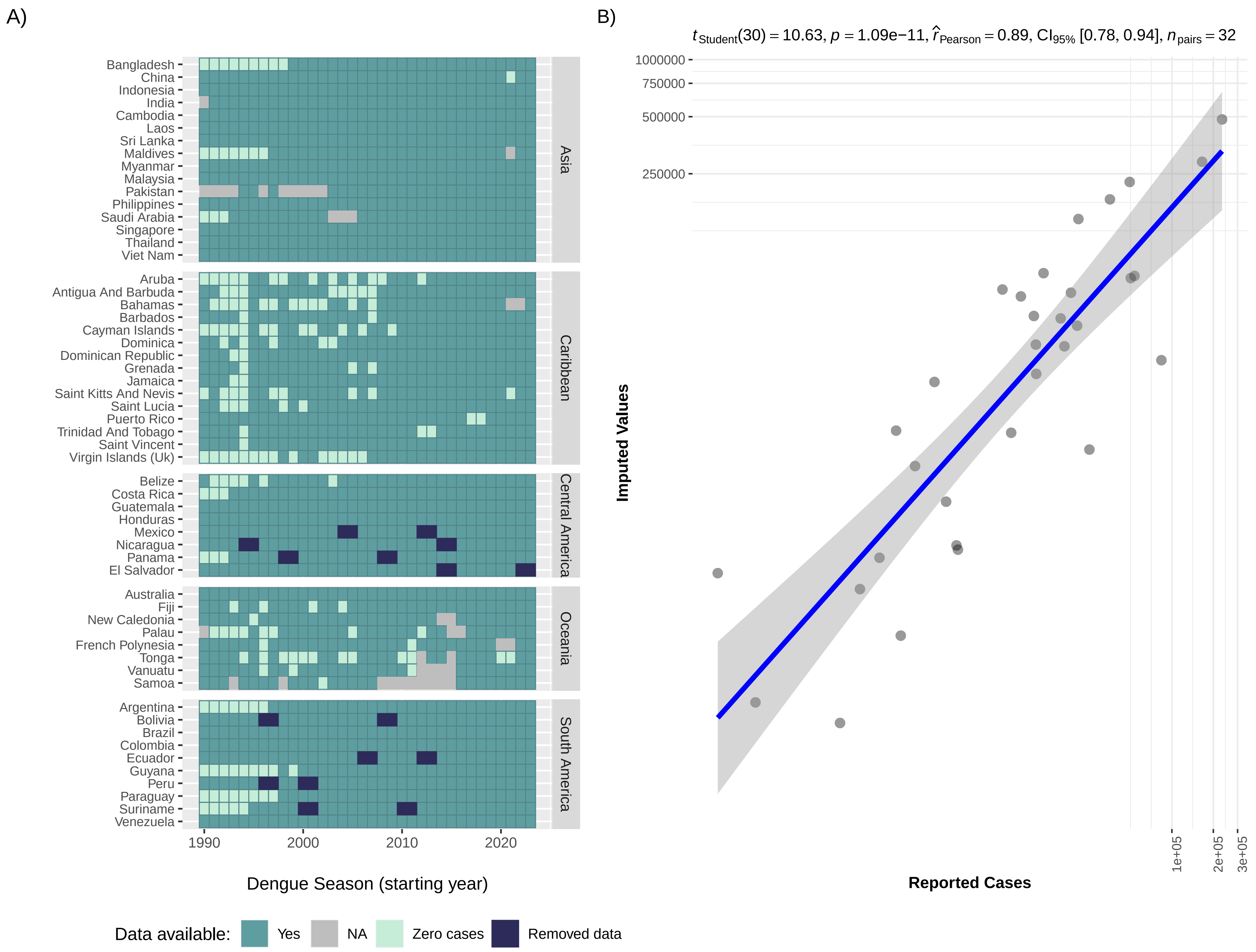

### Figure S9

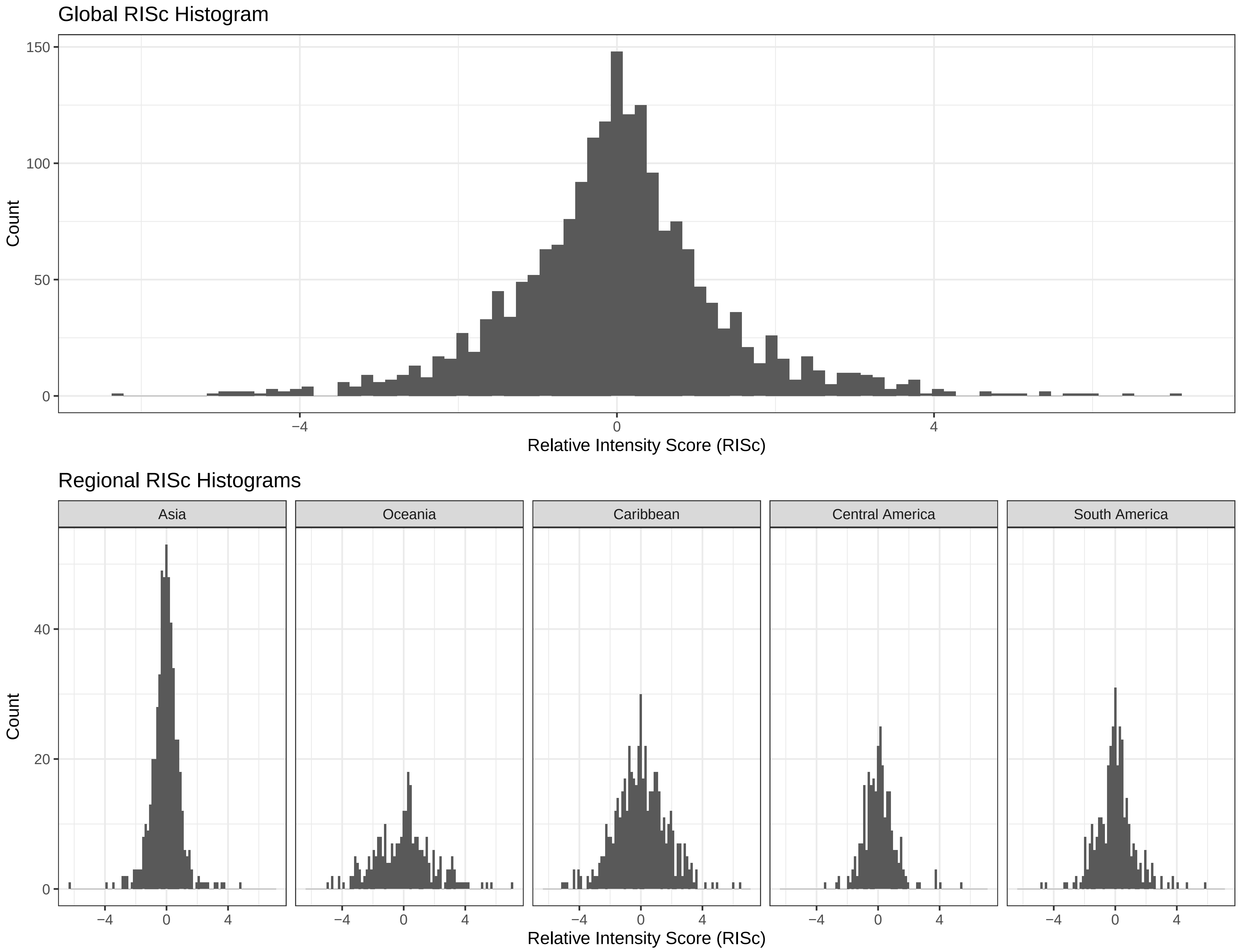

### Figure S10

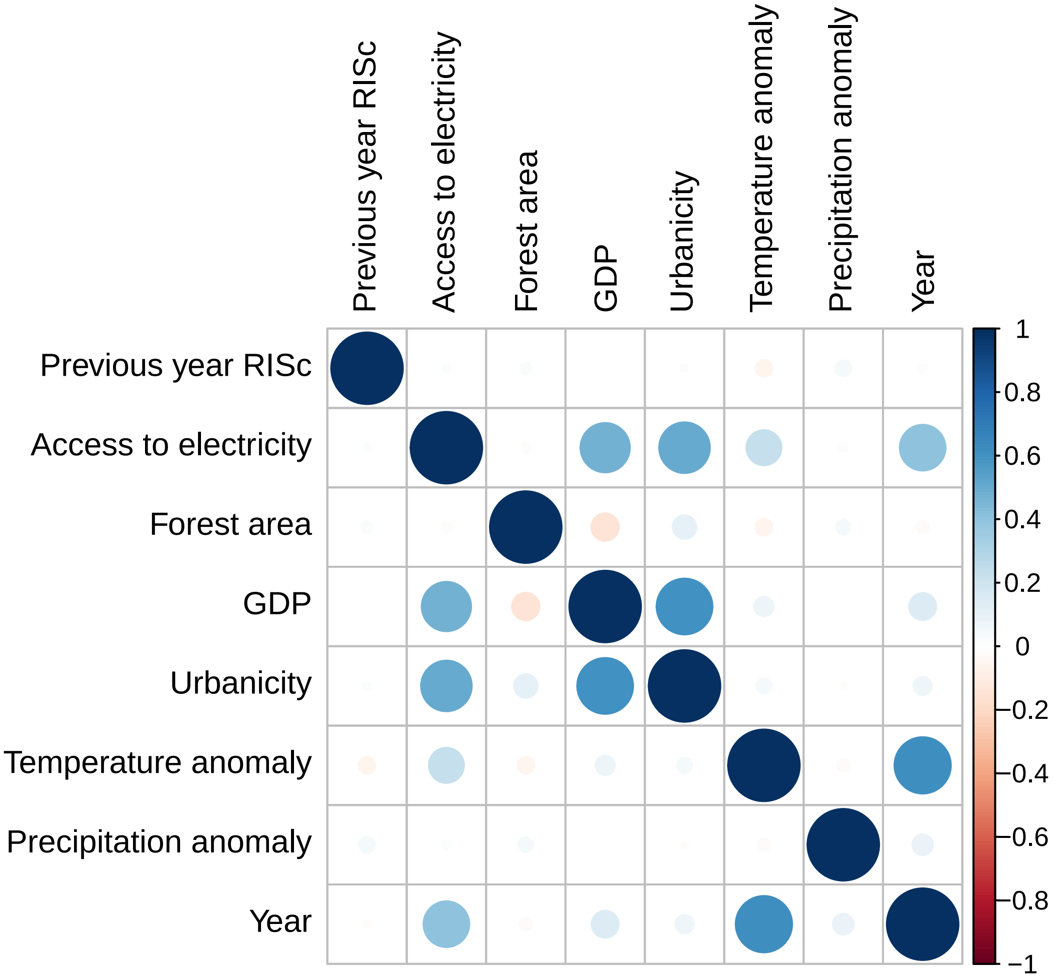

### Figure S11

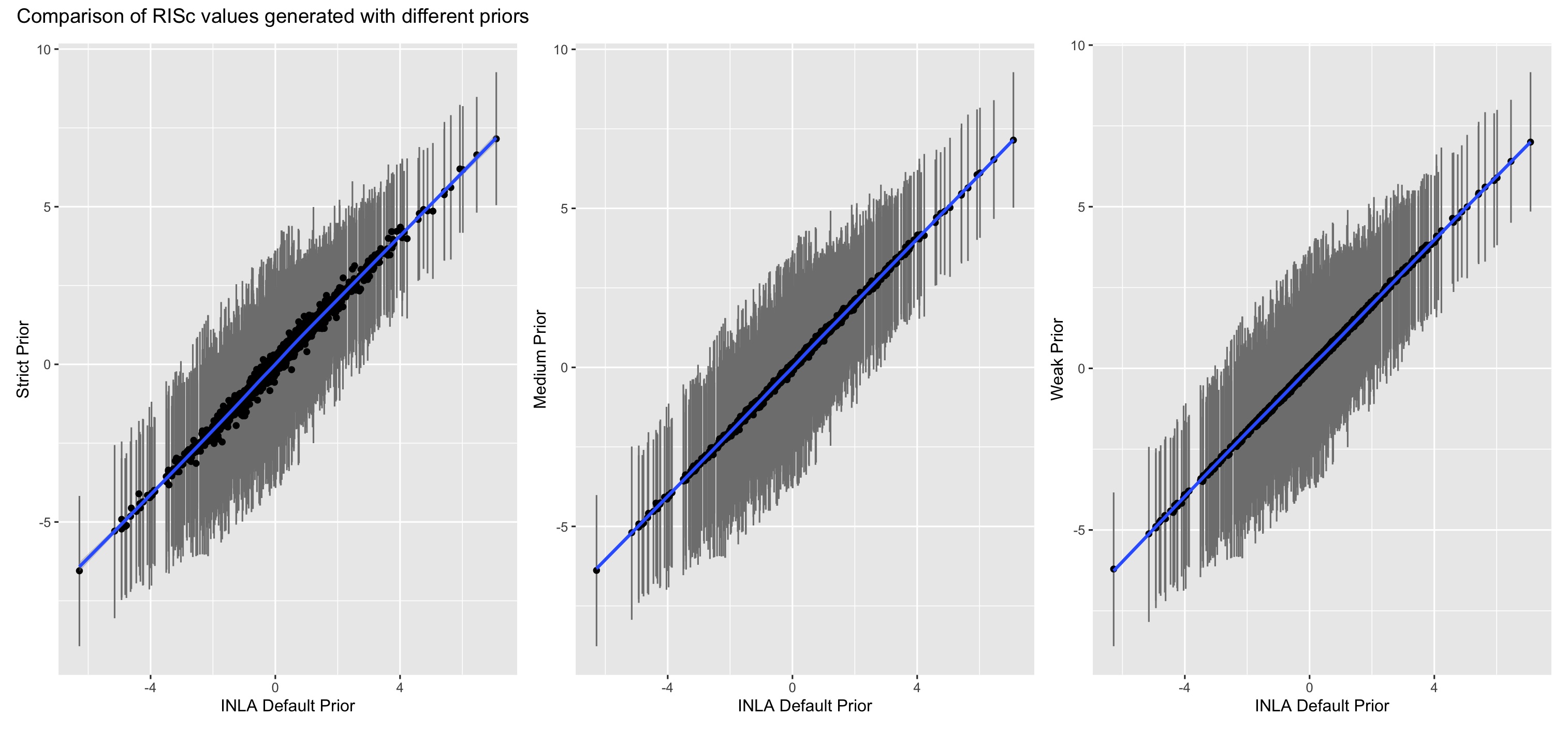

### Figure S12

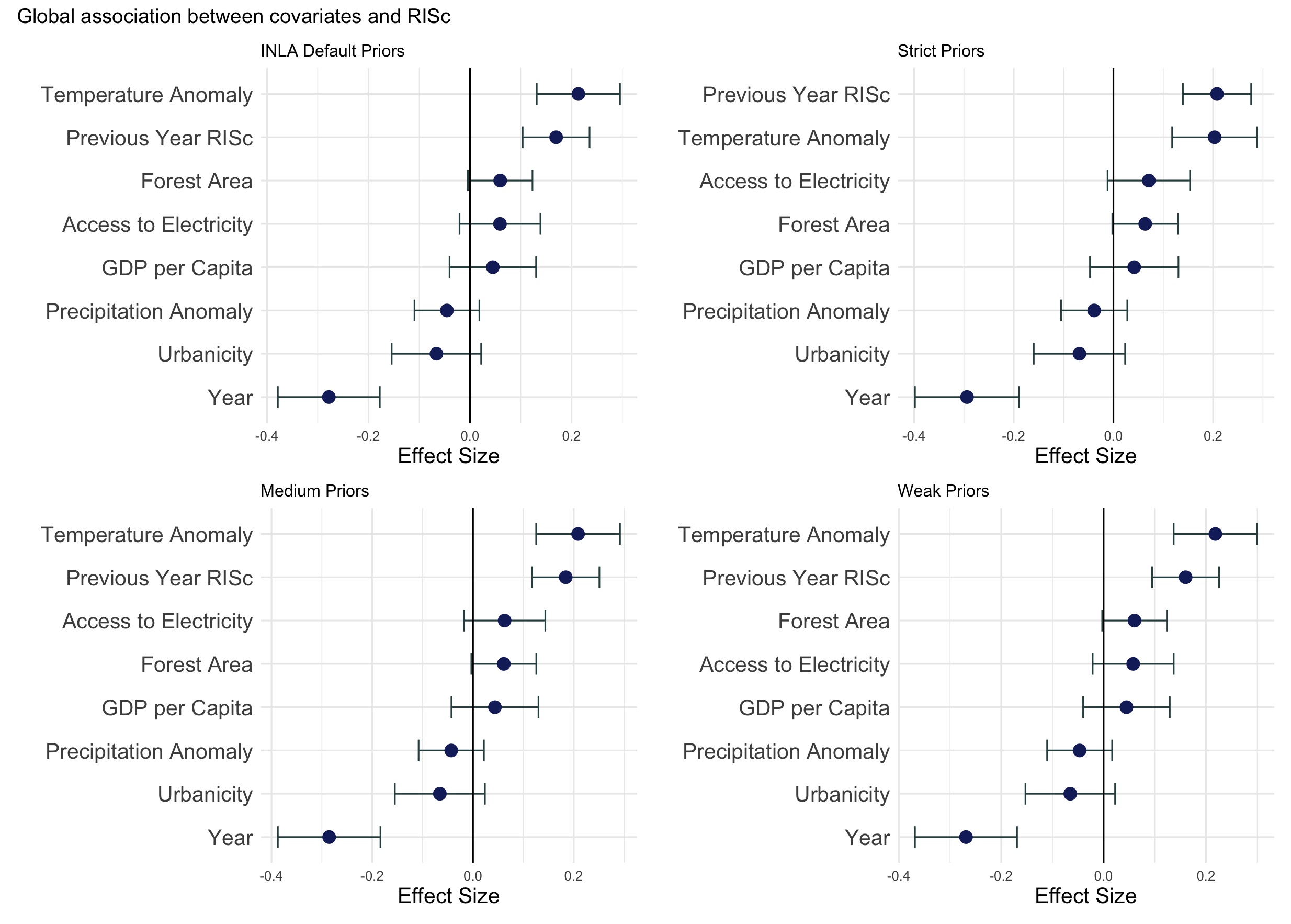
